# Longitudinal Trajectories of Child and Youth Mental Health Symptoms Across Distinct Phases of the COVID-19 Pandemic: A population-based study in Ontario, Canada

**DOI:** 10.64898/2026.04.02.26350051

**Authors:** Katholiki Georgiades, Yun–Ju Chen, Dylan Johnson, Ryan Miller, Li Wang, Amanda Sim, Emma Nolan, Nicole Dryburgh, Jordan Edwards, Seamus O’byrne, Ruth Repchuck, Katherine T. Cost, Laura Duncan, Meira Goldberg, Eric Duku, Peter Szatmari, Stelios Georgiades, Harriet L. MacMillan, Charlotte Waddell

**Author notes:** **Correspondence**: Katholiki (Kathy) Georgiades.

## Abstract

**Background:** Although an expansive body of evidence exists on children’s mental health during the COVID-19 pandemic, it is largely restricted to the early phases and lockdowns. This study examines longitudinal changes in child and youth mental health symptoms across two years of the COVID-19 pandemic, with data collection strategically timed to capture variability in pandemic restrictions.

**Methods:** A population-based longitudinal study of 1,261 children and youth aged 4–17 years followed prospectively from January 2021 to December 2022, with five waves of data collected in Ontario, Canada. Latent growth curve modelling was used to estimate trajectories of parent-reported mental health symptoms and identify baseline and time-varying covariates associated with variable trajectories.

**Findings:** Mental health symptoms were elevated and stable during lockdowns, followed by significant reductions as pandemic restrictions loosened, particularly for oppositional defiant and inattention/hyperactivity symptoms compared to internalizing symptoms. Children without pre-existing clinician diagnosed physical, mental or neurodevelopmental conditions and those not in lockdown at baseline demonstrated relative increases in mental health symptoms during lockdowns; and girls, compared to boys, demonstrated smaller reductions in internalizing symptoms as restrictions loosened. Concurrent and lagged associations between parental distress and children’s mental health symptoms varied across the pandemic.

**Interpretation:** Variation in symptom trajectories by mental health domain, gender, pandemic restrictions and pre-existing diagnosed conditions underscores the need for tailored, equity-informed pandemic planning and response. Policies designed to optimize the balance between the need to reduce viral community transmission whilst limiting pandemic lockdowns may mitigate adverse impacts on child and youth mental health.

**Funding:** Ontario Ministry of Health

## Introduction

During the COVID-19 pandemic, children and youth experienced major health, economic, and psychosocial stressors within their families and communities, alongside widespread school closures and reduced access to essential community services. Many of these stressors represent well-established risk factors for child and youth mental health, operating directly to influence mental health, or indirectly, by influencing intermediary factors such as parental mental health and interpersonal processes. Although there is a considerable body of evidence examining the mental health impacts of the COVID-19 pandemic on children and youth, systematic reviews have consistently documented substantial heterogeneity across studies and high risk of bias, making it difficult to draw robust conclusions (Ahmed et al., 2023; Ching et al., 2025; Geoffroy et al., 2024; Ludwig-Walz et al., 2022; Madigan et al., 2023; Newlove-Delgado et al., 2023; Sun et al., 2023). Common methodological limitations across studies include reliance on convenience sampling, cross-sectional designs, low response and retention, online data collection and limited representation of marginalized populations (Ahmed et al., 2023; Ching et al., 2025; Geoffroy et al., 2024; Ludwig-Walz et al., 2022; Madigan et al., 2023; Newlove-Delgado et al., 2023; Sun et al., 2023).

Existing studies have focused primarily on depression and anxiety in adolescent and young adult samples, with significant evidence gaps for younger children and other domains of mental health concerns, including externalizing and neurodevelopmental concerns (Geoffroy et al., 2024; Newlove-Delgado et al., 2023; Sun et al., 2023). Evidence is also primarily restricted to the first year of the pandemic and periods of lockdown, limiting understanding of how child and youth mental health changed in relation to subsequent easing and reimplementation of pandemic restrictions. These limitations underscore the ongoing need for high-quality, longitudinal, population-based data to support children and youth experiencing ongoing mental health difficulties and to inform policy responses for future pandemics and public health crises (Cortese et al., 2022; Demkowicz et al., 2021; Ford et al., 2024; Patten et al., 2021; Pierce et al., 2020).

The Canadian context provides an important opportunity to examine changes in child and youth mental health alongside evolving pandemic responses (Razak et al., 2022). Relative to other G10 countries, Canada implemented some of the most restrictive and prolonged public health measures during the first two years of the pandemic, including the second-longest duration of school closures (∼51 weeks; Razak et al., 2022). With successive waves of the COVID-19 pandemic, restrictions eased and tightened providing a unique context to examine the time-varying relationship between public health restrictions and changes in child and youth mental health symptoms over the course of the pandemic. Yet, evidence from Canada remains very limited (Sun et al., 2023; Newlove-Delgado et al., 2023; Miao et al., 2023; Geoffroy et al., 2024; Madigan et al., 2023).

Systematic reviews consistently highlight critical gaps in longitudinal, population-based evidence on child and youth mental health during the COVID-19 pandemic, particularly in Canada. In a review of 134 cohort studies comparing mental health outcomes before and during the pandemic, only 30 included children and adolescents (predominantly adolescents), almost all were conducted in the first year of the pandemic (n=28) and none were from Canada (Sun et al., 2023). Similarly, a review of the global evidence on mental health of children and young people, before versus during the pandemic, identified 51 studies, with only eight from Canada and zero rated as high quality (Newlove-Delgado et a., 2023). A review of Canadian studies on youth mental health during the COVID-19 pandemic also found that available evidence was primarily restricted to the first year of the pandemic (especially the first few months) and characterized by substantial heterogeneity and high risk of bias (Geoffroy et al., 2024).

Differences in COVID-19 related governmental responses between countries and within countries over time underscore the importance of examining changes in mental health in relation to evolving public health measures. The objectives of the present study were to estimate trajectories of parent-reported internalizing, oppositional defiant, and inattention/hyperactivity symptoms during distinct phases of the COVID-19 pandemic and identify baseline and time-varying covariates associated with variable trajectories. The study was designed to address several limitations of existing evidence by using a probability-based sample of the general population of children and youth aged 4–17 years in Ontario, Canada and longitudinal data collection timed to maximize variability in public health restrictions across five distinct phases of the pandemic from January 2021 to December 2022. At baseline, the province of Ontario took a regional approach to public health restrictions, resulting in variability across the sample with respect to lockdown status (i.e., some children were in lockdown whereas others were not). At the subsequent wave of data collection, the province of Ontario was in complete lockdown. This study design provides a unique opportunity to compare children and youth not in lockdown versus those in lockdown at baseline and the transition to lockdown status at Time 2, and to assess mental health-related changes associated with the transition from lockdown to reopening phases with varying levels of restrictions. Reporting for the present study follows the Strengthening the Reporting of Observational Studies in Epidemiology (STROBE) guidelines for cohort studies (von Elm et al., 2007).

## METHODS

### Study Design and Participants

The COVID-19 **On**tario **P**rovincial **A**ssessment and **T**racking of Child and Family **H**ealth Study (ONPATH) is a general population-based longitudinal study designed to examine changes in mental health symptoms and access to care among children and youth aged 4-17 years and their families during the COVID-19 pandemic living in Ontario, Canada. The sample was drawn from the Prob*it* panel (Probit, 2024), a probabilistic survey research panel available in Canada designed to reflect the demographic and socio-economic characteristics of the Canadian population. The sampling frame included all Prob*it* panel members with at least one child aged 4–17 years living in their household in Ontario as of January 10, 2021 (*n*=4,242) and was stratified by gender, level of education and region. Simple random samples were drawn from each stratum to reflect the distribution according to the 2016 Canadian Census (Statistics Canada, 2024a). A total of 1,261 panel members were eligible and agreed to participate (29·7% response rate). Comparisons against the 2016 Canadian Census revealed under-representation of households in the Greater Toronto Area and Central Regions, children aged 4–8 years and parents with a high school level of education. Sample weights were developed by deriving a weighting factor for each participant that adjusts the sample distribution to most closely match the target population of Ontario households with children aged 4–17 years. Weighted baseline demographic and socio-economic sample characteristics are presented in Table 1. For comparative purposes, weighted sample characteristics of the Ontario Child Health Study (OCHS; Boyle et al., 2019a), a mental health epidemiologic survey of a representative sample of 10,802 children and youth aged 4–17 years in Ontario in 2014–2015 conducted by Statistics Canada, are also presented in Table 1. Sample descriptives stratified by age group and gender are included in Appendix A.

**Table 1.** Baseline Sample Characteristics, valid n (%)

|  | **ONPATH**<br>**(n=1,261)** | **OCHS**<br>**(n=10,802)** |
| --- | --- | --- |
| **Child Gender** |  |  |
| Boy | 627 (50.9) | 5541 (51.3) |
| Girl | 611 (48.1) | 5261 (48.7) |
| Other gender | 13 (1.0) | .. |
| **Child Age, Mean (SD)** | 11.19 (5.39) | 10.63 (4.07) |
| **Schooling Modality** |  |  |
| Full-time in-person | 408 (35.9) | .. |
| Blended | 249 (17.5) | .. |
| Full-time online | 553 (46.7) | .. |
| **Pandemic Lockdown Status at Baseline** |  |  |
| Lockdown | 1007 (80.5) | .. |
| Not Lockdown | 230 (19.6) | .. |
| **COVID-19 Case Rate per 100 000, Mean (SD)** | 8.30 (7.01) | .. |
| **Pre-existing Clinician Diagnosed Physical, Mental or Neurodevelopmental Condition** |  |  |
| Yes | 469 (35.3) | .. |
| No | 781 (64.7) | .. |
| **Parental Distress, Mean (SD)** | 4.95 (6.01) | 3.10 (3.59) |
| **Parent-Reported Child and Youth Mental Health Symptoms** |  |  |
| **Total Raw Score, Mean (SD)** |  |  |
| Internalizing Symptoms | 3.83 (4.95) | 1.65 (2.50) |
| Oppositional Defiant Symptoms | 3.65 (3.66) | 1.60 (2.07) |
| Inattention/Hyperactivity Symptoms | 4.72 (4.74) | 2.16 (2.77) |
| **Moderated Non-Linear Factor Analysis Score (MNLFA), Mean (SD)** |  |  |
| Internalizing Symptoms | -0.14 (1.07) | .. |
| Oppositional Defiant Symptoms | 0.17 (.98) | .. |
| Inattention/Hyperactivity Symptoms | 0.24 (1.03) | .. |
| **Parent Gender** |  |  |
| Man | 704 (57.7) | 1361 (12.6) |
| Woman | 551 (42.1) | 9441 (87.4) |
| Other gender | 2 (0.2) | .. |
| **Parent Age, Mean (SD)** | 47.26 (10.38) | 41.88 (7.11) |
| **Parent Racialized Identity** |  |  |
| White | 1015 (81.6) | 6762 (62.6) |
| Asian | 106 (6.4) | 2550 (23.6) |
| Black | 52 (4.7) | 540 (5.0) |
| Other/Mixed | 86 (7.3) | 951 (8.8) |
| **Low Income Household** |  |  |
| Low Income | 125 (16.6) | 1901 (17.6) |
| Not Low Income | 922 (83.5) | 8901 (82.4) |
| **Number of Biological Parents in Family** |  |  |
| Two biological parents | 992 (78.1) | 7723 (71.5) |
| One or no biological Parents | 263 (21.9) | 3079 (28.5) |
| **Parent Education** |  |  |
| Highschool or less | 92 (29.7) | 1383 (12.8) |
| Post-secondary diploma or certificate less than Bachelor's degree | 344 (33.4) | 4461 (41.3) |
| University degree, Bachelor's or above | 820 (36.9) | 4958 (45.9) |
| **Urban-rural Residency** |  |  |
| Large urban centre | 891 (69.2) | 7713 (71.4) |
| Small-medium centre | 182 (15.1) | 1750 (16.2) |
| Rural area | 159 (15.7) | 1339 (12.4) |

.. = data not collected in OCHS, n's unweighted, percentages are weighted, MNLFA = moderated non-linear factor analysis

Baseline data were collected January 13–February 24, 2021. Respondents who participated at baseline were invited to complete four, follow-up waves occurring every 4–6 months and/or timed to maximize variability in the stringency of societal restrictions imposed to limit the spread of SARS-CoV-2 infection. Data collection dates and retention rates were: Time 2, May 7–June 15, 2021, 76·0% (*n* = 959); Time 3, October 13–November 25, 2021, 66·1% (*n =* 833); Time 4, February 18–March 23, 2022, 61·8% (*n =* 779); Time 5, October 20–December 6, 2022, 57·0% (*n* = 719). Patterns of nonresponse and baseline characteristics of respondents at each wave are presented in Appendix B.

Figure 1 presents provincial data for Ontario’s COVID-19 Stringency Index (Hale et al., 2021), COVID-19 case rate per 100,000, and status of school closures and modifications in relation to each wave of data collection. At baseline, the province of Ontario took a regional approach to restrictions, with Public Health Units (PHUs) determining the nature and severity of nonpharmacologic public health measures. At Time 2, Ontario was in a state of complete lockdown with no variability across PHUs and all publicly-funded schools closed to in-person learning. Times 3 and 4 occurred during periods of re-opening with public health restrictions gradually lifting and schools open to full-time, in-person learning but with significant modifications. There was a brief intervening period between Times 3 and 4, owing to the Omicron variant, where restrictions increased and schools were closed to in-person learning. At Time 5, almost all restrictions were lifted with those remaining focused on protecting the most vulnerable. Schools were open to in-person learning with no modifications and the Oxford Stringency Index was at its lowest value throughout the study.

**Figure 1.**
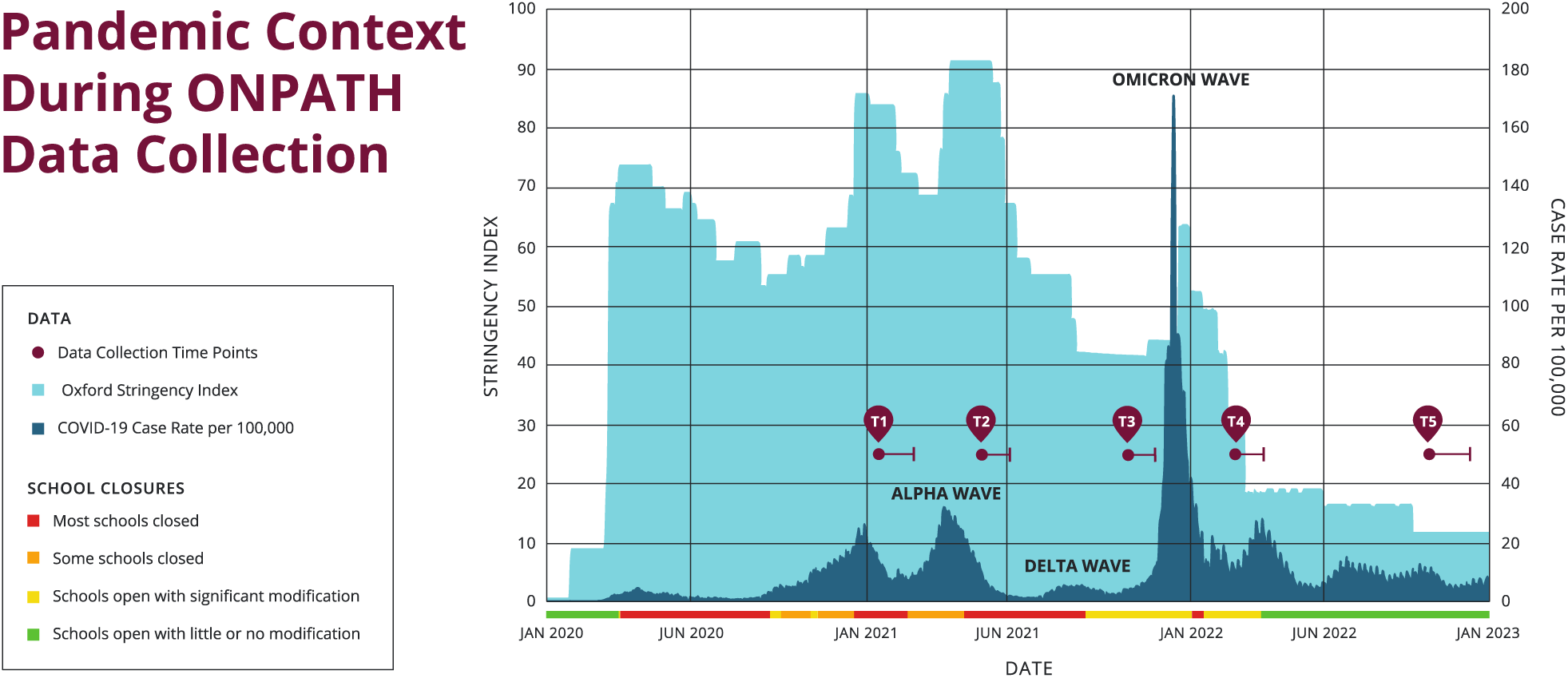
Pandemic context during ONPATH data collection. Figure 1 illustrates the state of the pandemic and concurrent public health measures meant to combat COVID-19 over the course of the ONPATH study. The graph shows the state of the pandemic on a provincial level. On the x-axis is the date from January 1, 2020, to January 1, 2023, covering from the start of the COVID-19 pandemic in Ontario through the ONPATH timepoints. The maroon timepoints are labelled T1-T5 with the solid point indicating the start of data collection for that timepoint, and the bar extending outward indicating the time until the last point of data was collected. The light blue shading and left y-axis indicates the Oxford stringency index, a composite measure of various public health restrictions such as school closures, public gathering limits, etc. The dark blue shading and right y-axis shows the COVID-19 case rate per 100 000 in Ontario and labels are given for the major variants of COVID-19 present during the dates shown, e.g. alpha wave. Finally, to highlight the importance of school closures, the colour of the x-axis of the graph reflects the school closure item of the Oxford stringency measures, with green representing little to no restrictions on schools, yellow indicating schools being open with significant modification e.g. social distancing, masking etc., orange represents the closure of some schools or recommendations to close, whereas red represents the mandatory closure of most schools.

### Procedures and Outcomes

Procedures remained the same across all waves of data collection and study materials were available in English and French. Invitations to participate were sent via e-mail, with follow-up telephone calls conducted by trained, bilingual interviewers to reach those who did not respond to email invitations. Invitations included a brief description of the study, confirmation of eligibility and consent to participate (online form or documented verbal consent). At each wave of data collection, participants were entered into a lottery draw for three prizes ($200 each). The Hamilton Integrated Research Ethics Board granted ethics approval (HiREB Project #: 10855).

Participants were given the option to complete the surveys online or by telephone at each wave of data collection (See Appendix A for these distributions). Participants with more than one eligible child living in their household were asked to select and consistently report on the child who had their most recent birthday at the time of the baseline survey or who was the last born in the case of twins or triplets. Participants whose selected child was 11 years of age or older were asked for consent to contact and administer a related, online youth survey.

#### Measures

##### Choice of primary measure

The primary outcome measure is an abbreviated version of the Ontario Child Health Study Emotional Behavioural Scales (OCHS-EBS, Duncan et al. 2019). The OCHS-EBS is a 52-item measure that includes dimensional symptom ratings for seven *DSM-5* disorders assessed by parents of children aged 4–17 years, including major depression, generalized anxiety, separation anxiety, social anxiety, conduct, oppositional defiant and attention deficit hyperactivity disorders. With the exception of conduct disorder, internal consistency and test-retest reliabilities of the parent scales are ≥ 0·70 (for a detailed description of the psychometric properties of the scales see Duncan et al., 2019 and Boyle et al., 2019b. The OCHS-EBS is freely available in English and French and has a long history of use in general population studies of child and youth health in Canada dating back to 1983 with the original scales (Offord et al., 1989; Boyle et al., 1993).

To address ONPATH’s research objectives, 29 OCHS-EBS items measuring the following disorders were selected for psychometric evaluation: major depression (nine items), generalized anxiety (six items), oppositional defiant (six items) and attention deficit/hyperactivity (eight items). Given low endorsement of conduct disorder symptoms in general population studies (Duncan et al., 2019; Boyle et al., 2019b), coupled with more limited evidence of reliability (Duncan et al., 2025; Duncan et al., 2019; Boyle et al., 2019b), this condition was excluded. Separation and social anxiety were also excluded given pandemic restrictions and more limited opportunities for socialization outside the home.

Psychometric analyses of the 29 selected items were conducted using data from the OCHS (Boyle et al., 2019a; see Appendix C for a detailed description). Exploratory and confirmatory factor analyses supported a three-factor model corresponding to internalizing, oppositional defiant and inattention/hyperactivity symptoms. A three-dimensional, two-parameter, item response theory model informed item selection, resulting in a 21-item measure including: eight internalizing, six oppositional defiant and seven inattention/hyperactivity items. Internal consistency (Cronbach’s α) of each mental health domain exceeded 0·9 and test-retest reliabilities (Pearson’s *r*) exceeded 0·7. Strong evidence for both internal and external convergent and discriminant validity (assessed against the Mini International Neuropsychiatric Interview for Children and Adolescents – MINIKID; Sheehan et al., 2010) was also found (see Appendix C).

This 21-item OCHS-EBS abbreviated measure was administered to parents at each wave of data collection. Parents were asked to rate how well each individual symptom described their child in the past week, from 0 = never or not true, 1 = sometimes or somewhat true, 2 = often or very true.

Moderated nonlinear factor analyses (MNLFA) were used to derive outcome scores for the primary longitudinal analyses using the ONPATH longitudinal sample (n=1,261). Derived factor scores that adjust for non-invariance across time and by select participant characteristics, yielding a standardized normal distribution (M=0, SD=1), were used as the primary outcome measures. Factor scores were derived separately for each mental health domain—internalizing, oppositional defiant and inattention/hyperactivity symptoms—with higher scores indicating elevated symptom levels relative to the sample mean. Detailed information on the MNLFA analyses, results and score derivation of the 21-item OCHS-EBS abbreviated measure are available in Appendix D.

To assess the clinical meaning of change, Cohen’s *d* for repeated measures was calculated to index standardized mean change in MNLFA-derived scores estimated from unconditional latent growth models for each mental health domain across the study period (Lakens, 2013). This provides an interpretable metric of the practical significance of average population-level change over time. Conventional benchmarks were used as descriptive guidelines for interpreting magnitude (small ≈ 0·2, medium ≈ 0·5, large ≈ 0·8).

##### Covariates

Sociodemographic and economic variables included child’s age (in years), child’s gender (0 = girl, 1 = boy), parent gender (0 = woman, 1 = man), number of biological parents in the family (0 = one or no biological parent, 1 = two biological parents), household income below the low-income measure (0 = not low income, 1 = low income; based on the 2019 before-tax cutoffs adjusting for inflation; Statistics Canada, 2024b; Bank of Canada, 2024), parent’s highest education level (high school or less = 0, post-secondary below a bachelor’s = 1, and bachelor’s or higher = 2) and urban-rural residency based on linking participants’ forward sortation number to their region of residence (0 = large urban, 1 = small-medium urban, and 2 = rural) based on population density and size in the 2016 census (Statistics Canada, 2024a).

Parent’s racialized identity was assessed by asking respondents to mark all of the racial or cultural groups they belonged to, adapted from Statistics Canada’s population group of person standard (Statistics Canada 2021; White, South Asian, East Asian, Southeast Asian, West Asian or Arab, Black African, Black Caribbean, Black Canadian or American, Latin American or Central American or South American, or Another group). Indigenous identity was assessed by asking whether a parent identified as First Nations, Métis or Inuk. The following categories were collapsed for the purposes of descriptive statistics: White (non-racialized, non-indigenous), Asian (South Asian, East Asian, Southeast Asian, West Asian or Arab), Black (Black African, Black Caribbean, Black Canadian or American), or Other/Mixed (Latin American or Central American or South American, First Nations, Métis, or Inuk, or Other). According to Statistics Canada (2022) and Canadian Institute for Health Information (2022) guidelines, if two options were selected and one was White, the parent was classified as belonging to the racialized identity group, otherwise if two or more racialized identities were selected, they were coded as Other/Mixed. For statistical modeling purposes, the racialized identity variable was binary coded, *1,* for non-racialized, non-indigenous identity (i.e., White), and *0*, for all racialized, other/mixed identities.

Presence of a pre-existing clinician diagnosed physical, mental or neurodevelopmental condition at baseline was assessed by asking parents to report on long-term conditions that their child might have that are expected to last, or have already lasted, six months or more that have been diagnosed by a health professional. Parents could report the presence or absence of a condition in three categories: physical health; learning or communication disorder or disability; or a mental health or developmental disorder. A positive response to at least one of these categories resulted in being classified as having a pre-existing condition (1), otherwise the child was classified as not having a pre-existing condition (0). Parents completed the Kessler-6 at each wave of data collection to assess levels of parental distress in the past 30 days (Kessler et al., 2002). The K-6 includes questions about how often an individual has felt nervous, hopeless, restless or fidgety, so depressed that nothing could cheer them up, that everything was an effort, or worthless. Response options were: none of the time = 0, a little of the time = 1, some of the time = 2, most of the time = 3, all of the time = 4 (Kessler et al. 2002). Items were summed to derive an overall parental distress score, pro-rating up to 25% missing items. Pro-ration was conducted by taking the average score of completed items and multiplying by six. Cronbach’s alpha for the K-6 was 0·85 at baseline, indicating good internal consistency.

COVID-19 related characteristics included whether participants lived in an area with lockdown measures in effect at the time of their baseline survey completion ascertained by linking participants forward sortation number with their public health unit and coded as 1 = lockdown, 0 = non-lockdown (Ontario Data Catalogue, 2021). This measure was only derived at baseline due to lockdown policy becoming province-wide after baseline. The daily COVID-19 case rate per 100,000 in participants’ public health units on the date of each survey completion were obtained through this linkage (Public Health Ontario, 2024). Learning modality for children and youth enrolled in school was asked of parents at each timepoint by asking: “How is [child name] being educated right now?” (0 = full-time in person, 1 = hybrid, 2 = full-time remote).

### Statistical analysis

We first estimated a series of unconditional univariate latent growth curve models (LGCM) with various functional forms (linear, quadratic, and piecewise) to determine the solution that best describes our data based on several model fit indices, with comparative Fit Index (CFI)/Tucker–Lewis Index (TLI) ≥·95 and the root-mean-square error of approximation (RMSEA) ≤·05 indicating good fit (Hu & Bentler, 1999). For piecewise LGCMs, modification indices and visual inspection of estimated trajectories were used to detect potential turning points or knots (Kwok, Luo, & West, 2010).

After determining the optimal functional form for each mental health domain based on model fit statistics and theoretical considerations, we added time-invariant covariates (TICs) collected at baseline - child’s age and gender, parent’s gender and racialized identity, low-income household, Public Health Unit (PHU) lockdown status, pre-existing condition, and family structure - predicting the latent growth parameters. We then added time-varying covariates (TVCs): parental distress, school modality, and COVID-19 case rate, allowing both contemporaneous and lagged effects. Additionally, the TVC effects of parental distress were reparametrized to be contemporary correlated and cross-lagged given the potential bidirectional relation between parental distress and children’s mental health (Bagner et al., 2013; May & Williams, 2022). The full model specification is shown in Appendix F (Figure F1). LGCMs were conducted separately for each mental health domain using sampling weights in Mplus Version 8.7.

The analytic sample included respondents with data from at least one timepoint (N=1,261). Full-information maximum likelihood estimation was used to handle missingness in observed data under a missing-at-random assumption. Parent’s education was also incorporated as an auxiliary variable given its association with attrition. Missing data were generally low with all variables having less than 3% missingness at the item level, except for low-income status (17% missing at baseline). The distribution of item non-response for study variables at each wave is presented in Appendix E. While associations between baseline characteristics and participant non-response at each follow-up did not reveal consistent patterns across waves, lockdown status at baseline, child’s age, number of biological parents at home, and parent’s education and racialized identity were associated with non-response at specific timepoints (see Appendix B).

## Results

### Unconditional latent growth trajectories of mental health symptoms

The piecewise models adequately described the observed data of the unconditional LGCM, indicating a turning point at T2 across all three mental health outcomes (CFI = 0·967–0·995, TLI = 0·958–0·994, RMSEA = 0·027–0·068; see Appendix F, Table F1). As shown in Figure 2, the latent growth parameter estimates indicated a stable trend from T1 to T2 across all mental health outcomes (Mean = -0·01 to 0·01, all SE = 0·03, *p* > 0·05; *d* = -0·01 to 0·01), followed by a significant decrease from T2 to T5 (Mean = -0·03 to -0·07, all SE = 0·01, *p* < 0·001; *d* = -0·18 to -0·39).

**Figure 2.**
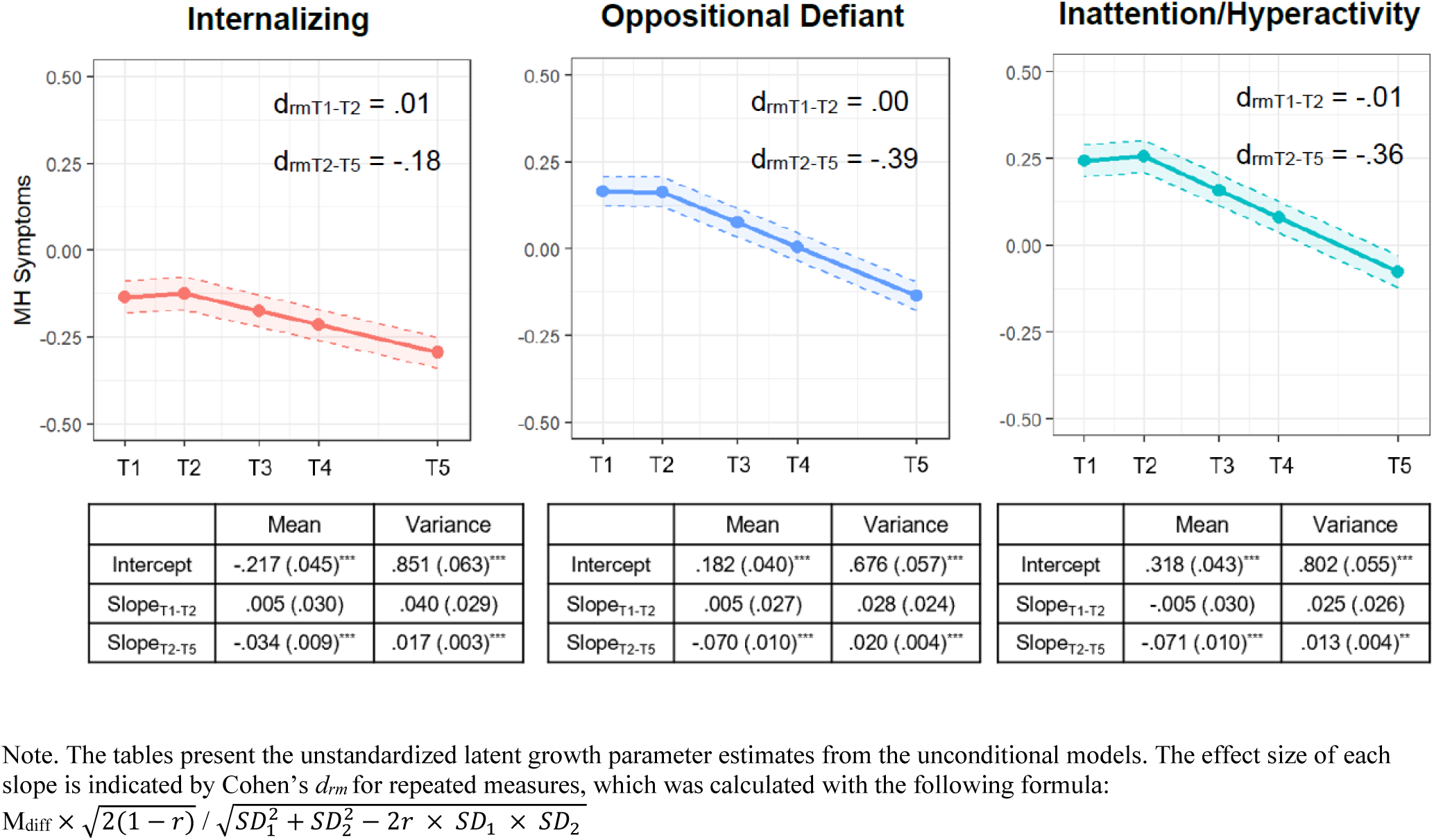
Weighted mean trajectories of mental health symptoms estimated by unconditional piecewise latent growth models

### Associations between time-invariant covariates (TICs) assessed at baseline and latent intercepts of mental health symptoms

At baseline, child’s older age was associated with higher levels of internalizing symptoms (β = 0·14, SE = 0·05, *p* < 0·001), but fewer oppositional defiant and inattention/hyperactivity symptoms (β = -0·19 & -0·40, SE = 0·05 & 0·04, both *p* < 0·001). Boys tended to have higher levels of inattention/hyperactivity symptoms (β = 0·14, SE = 0·04, *p* < 0·001), but fewer internalizing symptoms (β = -0·18, SE = 0·04, *p* < 0·001). Parents who identified as a woman and White were more likely to report higher levels of internalizing and inattention/hyperactivity symptoms (β = 0·09 to 0·17, all SE = 0·04 to 0·05, *p* < 0·05). Having a pre-existing condition was associated with higher scores across all mental health domains (β = 0·31 to 0·42, all SE = 0·04, *p* < 0·001). Lockdown status at baseline was associated with higher inattention/hyperactivity symptoms (β = 0·12, SE = 0·05, *p* = 0·011). Two-biological parent family was associated with lower oppositional defiant and inattention/hyperactivity symptoms (both β = -0·11, SE = 0·04 & 0·05, *p* < 0·05). All TIC associations with latent intercepts are described in Table 2.

**Table 2.** Weighted parameter estimates from full conditional piecewise latent growth curve model of mental health symptoms with time-invariant and time-varying covariates.

|  | Mental Health Symptom (MHS) Domains |  |  |
| --- | --- | --- | --- |
|  | Internalizing | Oppositional<br>Defiant | Inattention/<br>Hyperactivity |
| **Model fit** |  |  |  |
| $\chi^2$ (df) | 269.25 (140) | 280.22 (140) | 233.63 (140) |
| CFI/TLI | 0.97/0.93 | 0.97/0.92 | 0.98/0.95 |
| RMSEA [90% CI] | 0.03 [0.02, 0.03] | 0.03 [0.02, 0.03] | 0.02 [0.02, 0.03] |
| **Latent Growth Parameter Estimates** |  |  |  |
| **Mean (unstandardized)** |  |  |  |
| Intercept | -0.38 (0.17)* | -0.11 (0.18) | -0.40 (0.15)** |
| Slope $T_1-T_2$ | 0.08 (0.20) | 0.29 (0.24) | 0.02 (0.20) |
| Slope $T_2-T_5$ | 0.02 (0.06) | -0.02 (0.08) | 0.00 (0.07) |
| **R<sup>2</sup> (explained variance)** |  |  |  |
| Intercept | 0.32 | 0.15 | 0.34 |
| Slope $T_1-T_2$ | 0.27 | 0.32 | 0.21 |
| Slope $T_2-T_5$ | 0.06 | 0.04 | 0.04 |
| **Latent Growth Factor Correlation <sup>a</sup>** |  |  |  |
| Intercept ↔ Slope $T_2-T_5$ | -0.39*** | -0.27*** | -0.20** |
| **Time-Invariant Covariate Effects [standardized coefficients (SEs)]** |  |  |  |
| **Intercept** |  |  |  |
| Child age | 0.14 (0.05)** | -0.19 (0.05)*** | -0.40 (0.04)*** |
| Child gender (boy) | -0.18 (0.04)*** | -0.04 (0.04) | 0.14 (0.04)*** |
| Parent gender (man) | -0.17 (0.05)*** | -0.06 (0.05) | -0.09 (0.04)* |
| Parent racialized identity (white) | 0.13 (0.04)** | 0.13 (0.04)** | 0.11 (0.04)** |
| Low-income household | -0.03 (0.06) | -0.07 (0.05) | -0.02 (0.05) |
| Lockdown Status | 0.06 (0.05) | 0.10 (0.05)<sup>†</sup> | 0.12 (0.05)* |
| Pre-existing condition | 0.42 (0.04)*** | 0.31 (0.04)*** | 0.40 (0.04)*** |
| Two-parent family | -0.10 (0.05)<sup>†</sup> | -0.11 (0.05)* | -0.11 (0.04)* |
| **Slope $T_1-T_2$** |  |  |  |
| Child age | 0.15 (0.10) | -0.04 (0.14) | 0.06 (0.13) |
| Child gender (boy) | 0.05 (0.10) | -0.05 (0.14) | -0.06 (0.12) |
| Parent gender (man) | 0.34 (0.11)** | 0.11 (0.13) | 0.12 (0.13) |
| Parent racialized identity (white) | -0.12 (0.09) | -0.24 (0.15) | 0.03 (0.11) |
| Low-income household | 0.19 (0.12) | 0.22 (0.17) | 0.16 (0.13) |
| Lockdown Status | -0.23 (0.10)* | -0.31 (0.16)* | -0.35 (0.15)* |
| Pre-existing condition | -0.25 (0.11)* | -0.31 (0.15)* | -0.22 (0.13)<sup>†</sup> |
| Two-parent family | -0.06 (0.11) | 0.00 (0.13) | -0.00 (0.13) |
| **Slope $T_2-T_5$** |  |  |  |
| Child age | -0.10 (0.07) | 0.03 (0.08) | 0.07 (0.08) |
| Child gender (boy) | -0.17 (0.06)** | -0.05 (0.07) | -0.11 (0.08) |
| Parent gender (man) | 0.05 (0.07) | 0.01 (0.08) | 0.11 (0.08) |
| Parent racialized identity (white) | -0.11 (0.07) | -0.11 (0.08) | -0.03 (0.08) |
| Low-income household | 0.00 (0.06) | -0.04 (0.10) | -0.05 (0.08) |
| Lockdown Status | -0.01 (0.08) | -0.16 (0.07)* | -0.08 (0.08) |
| Pre-existing condition | 0.03 (0.07) | 0.02 (0.07) | 0.03 (0.07) |
| Two-parent family | 0.06 (0.07) | 0.02 (0.09) | 0.00 (0.08) |
| **Time-Varying Covariate Effects <sup>b</sup> [standardized coefficients (SEs)]** |  |  |  |
| **Parent Distress (PD)** |  |  |  |
| PD$_{T1}$ ↔ MHS$_{T1}$ | 0.30 (0.04)*** | 0.23 (0.04)*** | 0.23 (0.04)*** |
| $PD_{T2} \leftrightarrow MHS_{T2}$ | **0.23 (0.04)\*\*\*** | **0.16 (0.04)\*\*\*** | **0.16 (0.04)\*\*\*** |
| $PD_{T3} \leftrightarrow MHS_{T3}$ | **0.15 (0.04)\*\*\*** | **0.10 (0.04)\*\*** | **0.15 (0.03)\*\*\*** |
| $PD_{T4} \leftrightarrow MHS_{T4}$ | **0.19 (0.04)\*\*\*** | **0.13 (0.03)\*\*\*** | **0.15 (0.03)\*\*\*** |
| $PD_{T5} \leftrightarrow MHS_{T5}$ | **0.20 (0.08)\*** | 0.09 (0.08) | **0.28 (0.08)\*\*\*** |
| $PD_{T1} \rightarrow MHS_{T2}$ | **0.16 (0.03)\*\*\*** | **0.12 (0.03)\*\*\*** | **0.13 (0.03)\*\*\*** |
| $PD_{T2} \rightarrow MHS_{T3}$ | **0.09 (0.04)\*** | **0.10 (0.03)\*\*** | 0.01 (0.04) |
| $PD_{T3} \rightarrow MHS_{T4}$ | **0.13 (0.04)\*\*\*** | **0.10 (0.04)\*\*** | **0.10 (0.03)\*\*\*** |
| $PD_{T4} \rightarrow MHS_{T5}$ | **0.14 (0.03)\*\*\*** | **0.08 (0.04)\*** | 0.04 (0.03) |
| $MHS_{T1} \rightarrow PD_{T2}$ | **0.19 (0.03)\*\*\*** | **0.10 (0.03)\*\*** | **0.19 (0.03)\*\*\*** |
| $MHS_{T2} \rightarrow PD_{T3}$ | **0.17 (0.03)\*\*\*** | **0.13 (0.03)\*\*\*** | **0.17 (0.03)\*\*\*** |
| $MHS_{T3} \rightarrow PD_{T4}$ | **0.19 (0.04)\*\*\*** | **0.11 (0.04)\*\*** | **0.15 (0.03)\*\*\*** |
| $MHS_{T4} \rightarrow PD_{T5}$ | **0.16 (0.04)\*\*\*** | **0.12 (0.04)\*\*** | **0.13 (0.04)\*\*** |
| ***Schooling Mode (SM) [level of remoteness]*** |  |  |  |
| $SM_{T1} \rightarrow MHS_{T1}$ | 0.04 (0.03) | -0.02 (0.03) | 0.05 (0.03) |
| $SM_{T2} \rightarrow MHS_{T2}$ | 0.02 (0.02) | 0.01 (0.02) | **0.04 (0.02)\*** |
| $SM_{T3} \rightarrow MHS_{T3}$ | -0.02 (0.03) | -0.02 (0.03) | 0.02 (0.02) |
| $SM_{T4} \rightarrow MHS_{T4}$ | **0.12 (0.04)\*\*** | 0.05 (0.05) | 0.02 (0.05) |
| $SM_{T5} \rightarrow MHS_{T5}$ | 0.01 (0.03) | 0.01 (0.03) | 0.05 (0.03)<sup>†</sup> |
| $SM_{T1} \rightarrow MHS_{T2}$ | -0.01 (0.03) | **-0.08 (0.03)\*** | -0.02 (0.04) |
| $SM_{T2} \rightarrow MHS_{T3}$ | 0.02 (0.01) | -0.00 (0.02) | 0.00 (0.01) |
| $SM_{T3} \rightarrow MHS_{T4}$ | **-0.08 (0.04)\*** | -0.07 (0.05) | -0.02 (0.05) |
| $SM_{T4} \rightarrow MHS_{T5}$ | 0.03 (0.03) | 0.01 (0.04) | -0.02 (0.03) |
| ***Covid-19 Case Rate (CR)*** |  |  |  |
| $CR_{T1} \rightarrow MHS_{T1}$ | 0.02 (0.03) | **0.07 (0.03)\*** | -0.03 (0.03) |
| $CR_{T2} \rightarrow MHS_{T2}$ | 0.04 (0.03) | 0.03 (0.03) | 0.03 (0.03) |
| $CR_{T3} \rightarrow MHS_{T3}$ | 0.02 (0.03) | 0.04 (0.03) | -0.02 (0.02) |
| $CR_{T4} \rightarrow MHS_{T4}$ | -0.02 (0.03) | 0.03 (0.03) | 0.02 (0.03) |
| $CR_{T5} \rightarrow MHS_{T5}$ | -0.04 (0.03) | -0.02 (0.03) | -0.00 (0.03) |
| $CR_{T1} \rightarrow MHS_{T2}$ | -0.02 (0.03) | **0.07 (0.03)\*** | -0.01 (0.03) |
| $CR_{T2} \rightarrow MHS_{T3}$ | 0.03 (0.04) | 0.02 (0.04) | -0.03 (0.02) |
| $CR_{T3} \rightarrow MHS_{T4}$ | 0.01 (0.03) | 0.02 (0.03) | -0.04 (0.02) |
| $CR_{T4} \rightarrow MHS_{T5}$ | 0.04 (0.03) | 0.03 (0.04) | -0.02 (0.04) |

Note. \* $p < 0.05$ , \*\* $p < 0.01$ , \*\*\* $p < 0.001$ , <sup>†</sup> $p < 0.10$ .
<sup>a</sup>Covariances between the Slope<sub>T1-T2</sub> parameter and the other latent growth parameters (i.e., intercept and Slope<sub>T2-T5</sub>) were fixed to 0 for model identification purpose. <sup>b</sup>Covariances between TVCs were modeled but omitted here.

### Associations between TICs assessed at baseline and latent slopes of mental health symptoms

Lockdown status at baseline was associated with attenuated increases from T1 to T2 across all mental health symptom domains, compared to non-lockdown status (β = -0·23 to -0·35, SE = 0·11 to 0·16, both *p* < 0·05). Figure 3 presents the weighted mean mental health symptom trajectories from T1 to T2 by lockdown status at baseline. While there was a significant increase across all three mental health symptom domains from T1 to T2 for children who were not in lockdown at baseline and transitioned to lockdown status at T2 (Means = 0·12 to 0·17), symptom levels remained relatively stable for children who were in lockdown at baseline and remained in lockdown at T2 (Means = -0·05 to 0·02). Lockdown status at baseline was further associated with greater decreases in oppositional defiant symptoms from T2 to T5 (β = -0·16, SE = 0·07, *p* = 0·016). Having a pre-existing condition was associated with smaller increases in internalizing and oppositional defiant symptoms from T1 to T2 (β = -0·25 & -0·30, SE = 0·11 & 0·15, both *p* < 0·05). Children whose parent identified as a man demonstrated a tendency for greater increases in internalizing symptoms from T1 to T2 (β = 0·34, SE = 0·11, *p* = 0·001). Boys tended to have greater reductions in internalizing symptoms from T2 to T5 (β = -0·17, SE = 0·06, *p* = 0·010), compared to girls. All TIC associations with latent slopes are described in Table 2.

**Figure 3.**
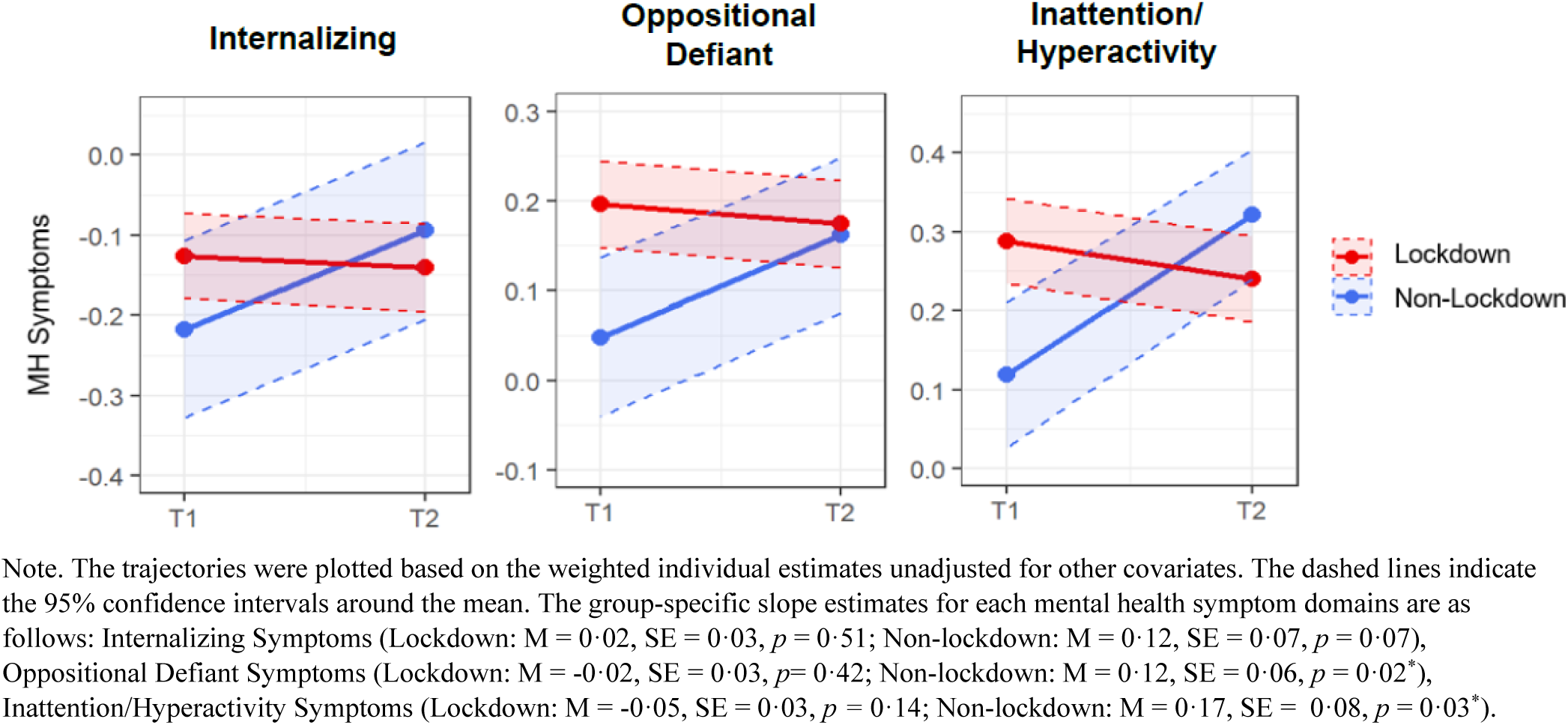
Weighted mean trajectories of mental health symptoms from T1 (baseline) to T2 by lockdown status at baseline.

### Effects of time-varying covariates (TVCs) on mental health symptoms

All TVC effects are presented in Table 2. Parent distress was overall positively associated with mental health symptoms from T1 to T5, except for oppositional defiant symptoms at T5 (Table 2). The concurrent correlations between parent distress and child mental health symptoms were stronger at T1 (*r* = 0·23 to 0·30, *p* <0·001) compared to T3 (*r* = 0·10 to 0·15, all *p* < 0·001), and then strengthened at T5 for internalizing and inattention/hyperactivity symptoms (*r* = 0·20 and 0·28, both *p* < 0·001).

Lagged effects of child mental health symptoms on parent distress remained relatively similar in magnitude from T1 to T5 (β = 0·10 to 0·19, SE = 0·03 to 0·04, all *p* < 0·01). The lagged effects in the opposite direction (parent distress → child mental health symptoms) remained relatively similar for internalizing and oppositional defiant symptoms from T1 to T5, with greater fluctuations evident for inattention/hyperactivity symptoms.

Higher internalizing symptoms at T4 were predicted by a higher level of school remote learning at T4 (β = 0·12, SE = 0·04, *p* = 0·008) and lower levels of remote learning at T3 (β = -0·08, SE = 0·04, *p* = 0·048). More oppositional defiant symptoms at T2 were associated with lower levels of remote learning at T1 (β = -0·08, SE = 0·03, *p* = 0·026). More inattention/hyperactivity symptoms at T2 were associated with concurrent remote schooling (β = 0·04, SE = 0·02, *p* = 0·042). Finally, higher oppositional defiant symptoms at T1 and T2 were associated with higher COVID-19 case rates at T1 (both β = 0·07, SE = 0·03, *p* < 0·05).

## Discussion

The present study examined longitudinal changes in child and youth mental health symptoms across different phases of the COVID-19 pandemic and the extent to which socio-demographic, socio-economic, pandemic, and health-related factors were related to these changes. Generally, mental health symptoms were elevated and stable during provincial lockdowns but decreased significantly as stringency measures were eased. The observed decreases in mental health symptoms during loosening of stringency measures were more pronounced for oppositional defiant and inattention/hyperactivity symptoms than for internalizing symptoms. Relative to those in lockdown at baseline, children who were not in lockdown demonstrated steeper increases in mental health symptoms from T1 to T2. Children with pre-existing clinician-diagnosed physical, mental, or neurodevelopmental conditions showed attenuated increases in internalizing and oppositional defiant symptoms from T1 to T2. Reductions in internalizing symptoms were attenuated for girls, compared to boys, from T2 to T5. Finally, parent distress and child mental health symptoms showed robust concurrent and lagged associations over time, with parent distress predicting subsequent child symptoms and child symptoms predicting subsequent parent distress across all time points.

Consistent with prior studies (Geoffroy et al., 2024), our findings align with evidence that child and youth mental health symptoms tend to be elevated during periods of heightened public health restrictions and subsequently decline as restrictions ease. For example, Geoffroy et al., (2024) reported increased depressive and anxiety symptoms during periods of greater pandemic-related disruption, followed by symptom improvement as restrictions were relaxed. Although the present study does not include a pre-pandemic comparison, symptoms in our sample were elevated and relatively stable during periods of peak restrictions and declined thereafter, suggesting a similar pattern of restriction-linked symptom elevation rather than a uniform or sustained deterioration over time. Importantly, these findings underscore that studies relying solely on assessments conducted during lockdown periods may overestimate the persistence of pandemic-related mental health impacts and misrepresent the full continuum of change as contextual conditions evolve.

While the pattern of longitudinal changes in mental health symptoms was mostly consistent, there are some notable differences that are important to acknowledge. The reductions in mental health symptoms observed as restrictions eased was less pronounced for internalizing symptoms, compared to oppositional defiant and inattention/hyperactivity symptoms. A potential explanation for this difference may relate to evidence suggesting that oppositional defiant and hyperactive/inattentive symptoms are particularly sensitive to contextual structure and environmental demands, including school routines and behavioral expectations (Ogundele & Ayyash, 2023). The return to in-person schooling may have facilitated symptom attenuation not only through access to formal behavioral interventions but also through the restoration of consistent structure, monitoring, and social regulation that may more strongly influence these behaviors than internalizing symptoms. In contrast, the attenuated symptom reductions observed for internalizing symptoms may reflect sustained disruptions in social connection, with evidence suggesting persistent feelings of loneliness following the pandemic (Einav & Margalit, 2023). Importantly, this pattern was particularly evident among girls, who showed attenuated decreases in internalizing symptoms from T2 to T5 relative to boys. This finding aligns with prior evidence documenting greater persistence of internalizing difficulties among girls during the pandemic (Geoffroy et al., 2024; Madigan et al., 2023). Although developmental trends, whereby internalizing symptoms tend to increase more steeply for girls, compared to boys, as they transition into adolescence cannot be ruled out, the observed gender differences over this relatively short time frame suggest that girls may have experienced a slower recovery from pandemic-related stressors, even as restrictions eased.

As lockdown procedures were regionally governed at baseline, there was variability in pandemic lockdown status across our sample, providing a naturalistic comparison group. Following baseline, lockdown procedures were governed provincially, resulting in the sample being exposed to the same regulations thereafter. At baseline, children and youth who lived in regions that were under pandemic lockdowns had significantly higher inattention/hyperactivity symptoms as compared to those not in lockdown. Between T1 and T2, children and youth who transitioned into lockdown experienced larger increases in mental health symptoms, whereas those who remained in lockdown demonstrated relative stability in mental health symptoms. Overall, this suggests that the transition into lockdown may represent an acute mental health risk factor, and that over time, some level of adaptation may occur, resulting in a stabilizing of mental health symptoms. Supporting this degree of adaptation, Geoffroy et al., (2024) observed that depressive symptoms worsened when transitioning from the pre-pandemic to active pandemic period (March 2020 to August 2020), but that depression symptoms were comparable to pre-pandemic levels over an extended period of the pandemic from September 2020 to June 2021.

Generally, both concurrent and lagged associations of parent distress and child mental health were observed, where parent distress predicted subsequent child mental health symptoms across all time points, as well as child mental health symptoms being associated with parent distress across all time points. This is consistent with literature showing the bidirectional association between parent and child mental health (Allmann et al., 2022; Pérez-Edgar et al., 2021), as well as pandemic literature which has shown that pandemic-related stressors have had a significant impact on the functioning of parents and their children (Racine et al., 2022; Thomson et al., 2023; Wade et al., 2021; Browne et al., 2021; Robertson et al., 2021). Some time-varying effects of schooling mode and pandemic case rates were observed for certain mental health domains; however, these effects were less consistent than those observed for parental distress, suggesting that family-level processes may have played a more central role in shaping child mental health during this period (Stracke et al., 2023).

The strengths of this study include its broad age range and the assessment of multiple child and youth mental health domains, including internalizing, oppositional defiant, and inattention/hyperactivity symptoms, alongside parent distress and its dynamic relationship with child mental health. The study employed a longitudinal design with data collection conducted via both telephone and internet modalities, enhancing accessibility and continuity of participation across the pandemic. Importantly, the extended duration of follow-up allowed the study to capture mental health trajectories across multiple pandemic phases, including the Alpha, Delta, and Omicron waves, as well as the period approaching the end of the pandemic, during which public health restrictions were progressively relaxed. A further strength is the use of regional and temporal variability in public health stringency across Ontario, enabling a naturalistic comparison of children and youth exposed to differing levels of lockdown conditions, an approach that remains relatively rare in the pandemic mental health literature. While the sampling frame was provincially representative and comparatively robust relative to many pandemic studies, it nonetheless shares limitations common to probability-based panel surveys (Si, Wagner, & Kessler, 2025). Despite reliance on parent-reported outcomes, the use of MNLFA scores represents a methodological strength, as this approach accounts for item-level differential functioning related to parent and child characteristics and ensures comparability of scores across time and respondents, thereby improving the accuracy of symptom measurement and mitigating reporting bias (Olino et al., 2021). Finally, the study incorporated multiple time-invariant and time-variant covariates into analyses, providing a more nuanced understanding of the temporal dynamics of mental health symptoms and individual, familial and socio-contextual correlates.

One limitation of this study is the absence of pre-pandemic mental health assessments. Data collection began approximately ten months after the World Health Organization declared COVID-19 a global pandemic, following a period marked by repeated provincial lockdowns, prolonged school closures, and substantial disruptions to daily routines between March 2020 and January 2021. Prior research suggests that the most pronounced changes in child and youth mental health occurred during the initial transition into the pandemic, therefore, the present study cannot directly characterize symptom changes during this early phase or disentangle their relation to trajectories observed later in the pandemic. More broadly, as with all observational COVID-19 research, causal inference is inherently limited. Leveraging regional and temporal variability in public health stringency provided us with a unique opportunity to examine pandemic-related impacts in the absence of feasible control conditions. Additional limitations include reliance on parent-reported outcomes, which may be subject to informant bias, particularly under conditions of elevated parent stress (Ford et al., 2024). Although the use of MNLFA scores helped mitigate some reporting bias, residual informant effects cannot be ruled out. Finally, the study was affected by both a modest initial response rate (∼30%) and attrition over time (∼43% by T5), raising the possibility of survivorship bias and an overestimation of mental health recovery (Czeisler et al., 2021). Consistent with this concern, respondents lost to follow-up were more likely to be single parents and have lower levels of education, and several sociodemographic groups were under-represented relative to the Ontario Child Health Study sampling frame, potentially limiting generalizability.

This study provides longitudinal evidence that child and youth mental health symptoms were elevated and remained relatively stable during periods of heightened pandemic restrictions, with symptom improvement observed as public health stringency measures eased. These patterns were not uniform across mental health symptom domains or subgroups: internalizing symptoms showed a more gradual decline compared to oppositional defiant and inattention/hyperactivity symptoms, particularly among girls, highlighting important gender disparities. Our findings further underscore the significance of contextual transitions and reveal a robust, bidirectional association between parent distress and child mental health symptoms, consistent with transactional models of family stress. Taken together, these results emphasize that pandemic-related impacts on child and family mental health unfold dynamically over time and may not be fully captured by assessments conducted during single phases of pandemic restrictions. Continued investment in longitudinal studies is essential to identify groups at risk for persistent difficulties, to inform timely and equitable mental health supports, and to strengthen preparedness for future public health crises.

## Data Availability

All data produced in the present study are available upon reasonable request to the authors.

