## Appendices for "Longitudinal Trajectories of Child and Youth Mental Health Symptoms Across Distinct Phases of the COVID-19 Pandemic: A population-based study in Ontario, Canada"

#### Supplementary Appendix

**Appendix A:** Baseline ONPATH Sample Descriptives Stratified by Gender and Age Group
**Appendix B**: Patterns of Participant Nonresponse at Follow-up Waves of Data Collection
**Appendix C**: Psychometric Analyses of Mental Health Symptom Measure
**Appendix D:** Derivation of Mental Health Symptom Domain Score based on Moderated Non-Linear Factor Analyses
**Appendix E:** Item Non-Response for Study Variables
**Appendix F**: Supplementary Results for the Primary Analysis

#### Appendix A: Baseline ONPATH Sample Descriptives Stratified by Gender and Age Group

**Table A1. ONPATH Sample descriptives stratified by gender and age group**

| | Total Sample (n=1,261) | Boy (n=627) | Girl (n=611) | 4–11 (n=563) | 12–17 (n=698) |
|---|---|---|---|---|---|
| **Child Gender, n (%)** | | | | | |
| Boy | 627 (50·9) | ·· | ·· | 281 (53·1) | 346 (47·9) |
| Girl | 611 (48·1) | ·· | ·· | 272 (46·3) | 339 (50·5) |
| Other gender | 13 (1·0) | ·· | ·· | 4 (0·6) | 9 (1·7) |
| **Child Age, Mean (SD)** | 11·19 (5·39) | 10·96 (4·11) | 11·09 (4·04) | 8·01 (2·37) | 15·03 (1·77) |
| **Schooling Modality, n (valid %)** | | | | | |
| Full-time in-person | 408 (35·9) | 193 (34·4) | 212 (38·1) | 234 (43·1) | 174 (26·8) |
| Blended | 249 (17·5) | 129 (19·5) | 114 (14·8) | 32 (7·9) | 217 (29·6) |
| Full-time online | 553 (46·7) | 285 (46·1) | 258 (47·1) | 265 (49·1) | 288 (43·7) |
| **Pandemic Lockdown Status at Baseline, n (valid %)** | | | | | |
| Lockdown [Lockdown, shutdown] | 1007 (80·5) | 496 (79·5) | 494 (81·5) | 445 (79·2) | 562 (82·1) |
| Not Lockdown [Prevent, Protest, Restrict, or Control] | 230 (19·6) | 117 (20·5) | 111 (18·5) | 108 (20·9) | 122 (17·9) |
| **COVID-19 Case Rate per 100 000, Mean (SD)** | 8·30 (7·01) | 8·14 (5·16) | 8·52 (5·33) | 8·40 (5·18) | 8·25 (5·44) |
| **Pre-Existing Clinician Diagnosed Physical, Mental or Neurodevelopmental Condition n (valid %)** | | | | | |
| Yes | 469 (37·5) | 245 (43·9) | 207 (34·0) | 169 (30·2) | 300 (43·4) |
| No | 781 (62·5) | 374 (56·1) | 402 (66·0) | 389 (69·8) | 392 (56·6) |
| **Parental Distress, Mean (SD)** | 4·95 (6·01) | 5·19 (4·67) | 4·75 (4·47) | 5·28 (4·79) | 4·56 (4·22) |
| **Parent-Reported Child and Youth Mental Health Symptoms** | | | | | |
| **Total Raw Score, Mean (SD)** | | | | | |
| Internalizing Symptoms | 3·83 (4·95) | 3·40 (3·65) | 4·08 (3·61) | 3·37 (3·40) | 4·36 (3·98) |
| Oppositional Defiant Symptoms | 3·65 (3·66) | 3·70 (2·80) | 3·61 (2·75) | 3·69 (2·69) | 3·66 (2·90) |
| Inattention/Hyperactivity Symptoms | 4·72 (4·74) | 5·31 (3·67) | 4·21 (3·29) | 5·38 (3·56) | 4·05 (3·53) |
| **Moderated Non-Linear Factor Analysis Score (MNLFA), Mean (SD)** | | | | | |
| Internalizing Symptoms | -0·14 (1·07) | -0·30 (1·04) | 0·00 (1·05) | -0·32 (1·00) | 0·01 (1·10) |
| Oppositional Defiant Symptoms | 0·17 (0·98) | 0·14 (0·96) | 0·02 (0·99) | 0·23 (0·90) | 0·11 (1·03) |
| Inattention/Hyperactivity Symptoms | 0·24 (1·03) | 0·38 (1·02) | 0·09 (1·01) | 0·51 (0·94) | 0·03 (1·05) |
| **Parent Gender, n (valid %)** | | | | | |
| Man | 704 (57·7) | 354 (60·2) | 340 (55·2) | 318 (58·1) | 386 (57·1) |
| Woman | 551 (42·1) | 271 (39·6) | 271 (44·8) | 241 (41·6) | 310 (42·9) |
| Other gender | 2 (0·2) | 1 (0·2) | 0 (0·0) | 2 (0·3) | 0 (0·0) |
| **Parent Age, Mean (SD)** | 47·26 (10·38) | 47·12 (7·55) | 47·16 (7·78) | 44·27 (6·42) | 50·95 (7·44) |
| **Parent Racialized Identity, n (valid %)** | | | | | |
| White | 1015 (81·6) | 499 (84·2) | 498 (78·5) | 440 (78·8) | 575 (85·2) |
| Asian [South Asian, East Asian, Southeast Asian, West Asian or Arab] | 92 (7·6) | 45 (5·6) | 45 (7·2) | 52 (7·8) | 40 (4·7) |
| Black [Black African, Black Caribbean, Black Canadian or American] | 48 (4·9) | 28 (5·1) | 20 (4·4) | 25 (5·5) | 23 (3·7) |
| Other/Mixed [Other, Latin American, Central American, South American, First Nations, Métis, Inuk] | 86 (5·9) | 45 (5·0) | 41 (9·9) | 40 (7·9) | 46 (6·4) |
| **Low Income Household, n (valid %)** | | | | | |
| Low Income | 125 (16·6) | 59 (14·0) | 60 (18·6) | 58 (16·7) | 67 (16·3) |
| Not Low Income | 922 (83·5) | 455 (86·0) | 456 (81·4) | 424 (83·3) | 498 (83·7) |
| **Number of Biological Parents in Family, n (valid %)** | | | | | |
| Two Biological Parents | 992 (78·1) | 496 (77·5) | 482 (79·0) | 469 (81·9) | 523 (73·0) |
| One or no biological Parents | 263 (21·9) | 128 (22·5) | 128 (21·0) | 92 (18·1) | 171 (27·0) |
| **Parent Education, n (valid %)** | | | | | |
| Highschool or less | 92 (29·7) | 55 (34·7) | 35 (25·1) | 50 (32·4) | 42 (26·1) |
| Post-secondary diploma or certificate less than Bachelor's degree | 344 (33·4) | 174 (32·5) | 165 (34·6) | 150 (32·4) | 194 (34·6) |
| University Degree, Bachelor's or above (2) | 820 (36·9) | 395 (32·8) | 410 (40·3) | 360 (35·2) | 460 (39·3) |
| **Urban-rural Residency, n (valid %)** | | | | | |
| Large urban centre | 891 (69·2) | 439 (67·5) | 436 (70·6) | 403 (70·2) | 488 (67·7) |
| Small-medium centre | 182 (15·1) | 86 (14·4) | 94 (16·1) | 71 (12·4) | 111 (18·7) |
| Rural area | 159 (15·7) | 86 (18·1) | 70 (13·3) | 76 (17·4) | 83 (13·5) |
| **Survey Mode, n (valid %)** | | | | | |
| Online | 796 (62·7) | 394 (61·2) | 386 (64·1) | 380 (65·5) | 416 (59·2) |
| Phone | 465 (37·3) | 233 (38·8) | 225 (35·9) | 183 (34·5) | 282 (40·8) |

n's unweighted, percentages are weighted.

#### Appendix B: Patterns of Participant Nonresponse at Follow-up Waves of Data Collection

All 1,261 participants who completed the baseline assessment were invited to complete the follow-up waves of data collection. Retention rates were 76·0% at Time 2 (*n*=959), 66·1% at Time 3 (*n*=833), 61·8% at Time 4 (*n*=779) and 57·0% (*n*=719). Patterns of nonresponse and baseline characteristics of participating respondents at each wave of data collection are presented in supplementary tables B1 and B2, respectively. Among all participants, 38·4% (n=484) completed all five waves, 20·3% (*n*=256) had one missing timepoint, 14·9% (*n*=188) missed two timepoints, 16·7% (*n*=210) missed three timepoints, and 9·8% (*n*=123) participated only at baseline (see table B1).

Baseline participant characteristics for each timepoint of data collection are presented in Table B2. Associations between participant non-response and baseline sample characteristics were examined with generalized estimating equations (GEE). Univariate GEE models were first conducted to identify significant correlates of non-response at each follow-up wave. The significant covariates were then included in a multivariate GEE model to obtain full-adjusted estimates. Lockdown status at baseline, child's age, number of biological parents in the home, and parental education and racialized identity were associated with participant non-response at specific follow-up timepoints (marked with asterisks in Table B2).

**Table B1. Patterns of Non-Response among the 1,261 Respondents to the ONPATH Study**

| | N | % |
|---|---|---|
| **Non-response at each timepoint** | | |
| Time 2 | 302 | 24·0 |
| Time 3 | 428 | 33·9 |
| Time 4 | 482 | 38·2 |
| Time 5 | 542 | 43·0 |
| **Missing number of timepoints** | | |
| All complete | 484 | 38·4 |
| 1 Time | 256 | 20·3 |
| 2 Times | 188 | 14·9 |
| 3 Times | 210 | 16·7 |
| 4 Times | 123 | 9·8 |
| **Breakdown of missing timepoints** | | |
| All complete | 484 | 38·4 |
| Time 2 Only | 46 | 3·6 |
| Time 3 Only | 48 | 3·8 |
| Time 4 Only | 45 | 3·6 |
| Time 5 Only | 117 | 9·3 |
| Times 2 & 3 Only | 11 | 0·9 |
| Times 2 & 4 Only | 13 | 1·0 |
| Times 2 & 5 Only | 23 | 1·8 |
| Times 3 & 4 Only | 39 | 3·1 |
| Times 3 & 5 Only | 30 | 2·4 |
| Times 4 & 5 Only | 72 | 5·7 |
| Times 2, 3, & 4 Only | 33 | 2·6 |
| Times 2, 3, & 5 Only | 20 | 1·6 |
| Times 2, 4, & 5 Only | 33 | 2·6 |
| Times 3, 4, & 5 Only | 124 | 9·8 |
| Times 2, 3, 4, & 5 | 123 | 9·8 |

All values are unweighted. All respondents participated at Baseline (Time 1).

**Table B2. Baseline Characteristics of Participating Respondents by Data Collection Time Point, valid n (%)**

| | Time 1 (n=1,261) | Time 2 (n=959) | Time 3 (n=833) | Time 4 (n=779) | Time 5 (n=719) |
|---|---|---|---|---|---|
| **Child Gender** | | | | | |
| Boy | 627 (50·1) | 477 (50·1) | 410 (49·6) | 379 (48·7) | 358 (50·2) |
| Girl | 611 (48·8) | 467 (49·0) | 409 (49·1) | 388 (49·8) | 350 (49·1) |
| Other gender | 13 (1·0) | 9 (0·9) | 7 (0·8) | 6 (0·8) | 5 (0·7) |
| **Child Age, Mean (SD)** | 12·12 (3·97) | 12·04 (3·96) | 11·93 (3·96)* | 12·06 (3·98) | 12·17 (3·95) |
| **Schooling Modality** | | | | | |
| Full-time in-person | 408 (33·7) | 302 (32·8) | 269 (33·6) | 245 (32·7) | 229 (32·9) |
| Blended | 249 (20·6) | 185 (20·1) | 167 (20·8) | 148 (19·8) | 142 (20·4) |
| Full-time online | 553 (45·7) | 434 (47·1) | 365 (45·6) | 356 (47·5) | 325 (46·7) |
| **Pandemic Lockdown Status at Baseline** | | | | | |
| Lockdown | 1007 (81·4) | 785 (82·8)* | 682 (82·9) | 647 (84·2)* | 592 (83·3) |
| Not Lockdown | 230 (18·6) | 163 (17·2) | 141 (17·1) | 121 (15·8) | 119 (16·7) |
| **COVID-19 Case Rate Per 100 000, Mean (SD)** | 7·93 (5·4) | 7·93 (5·4) | 7·80 (5·3) | 7·88 (5·4) | 7·97 (5·4) |
| **Pre-existing Clinician Diagnosed Physical, Mental or Neurodevelopmental Condition** | | | | | |
| Yes | 469 (37·5) | 358 (37·7) | 304 (36·9) | 289 (37·4) | 266 (37·6) |
| No | 781 (62·5) | 591 (62·3) | 520 (63·1) | 483 (62·6) | 441 (62·4) |
| **Parental Distress, Mean (SD)** | 4·83 (4·51) | 4·86 (4·48) | 4·69 (4·34) | 4·70 (4·40) | 4·80 (4·44) |
| **Parent-Reported Child and Youth Mental Health Symptoms** | | | | | |
| **Total Raw Score, Mean (SD)** | | | | | |
| Internalizing Symptoms | 4·04 (3·71) | 4·02 (3·66) | 4·09 (3·77) | 4·05 (3·73) | 3·98 (3·69) |
| Oppositional Defiant Symptoms | 3·67 (2·80) | 3·66 (2·78) | 3·65 (2·76) | 3·66 (2·82) | 3·59 (2·81) |
| Inattention/Hyperactivity Symptoms | 4·53 (3·51) | 4·56 (3·52) | 4·53 (3·46) | 4·53 (3·48) | 4·63 (3·52) |
| **Moderated Non-Linear Factor Analysis Score (MNLFA), Mean (SD)** | | | | | |
| Internalizing Symptoms | -0·14 (1·07) | -0·12 (1·04) | -0·21 (1·06) | -0·29 (1·07) | -0·29 (1·04) |
| Oppositional Defiant Symptoms | 0·17 (0·98) | 0·22 (0·99) | -0·00 (0·95) | -0·09 (0·97) | -0·12 (0·94) |
| Inattention/Hyperactivity Symptoms | 0·24 (1·03) | 0·36 (1·06) | 0·06 (0·98) | 0·00 (1·03) | -0·02 (1·02) |
| **Parent Gender** | | | | | |
| Man | 704 (56·0) | 543 (56·8) | 477 (57·5) | 450 (58·0) | 425 (59·3) |
| Woman | 551 (43·8) | 412 (43·1) | 353 (42·5) | 325 (41·9) | 291 (40·6) |
| Other gender | 2 (0·2) | 1 (0·1) | 0 (0·0) | 1 (0·1) | 1 (0·1) |
| **Parent Racialized Identity** | | | | | |
| White | 1015 (81·8) | 782 (81·5) | 678 (81·4) | 642 (83·7)* | 587 (82·9) |
| Asian | 106 (7·4) | 71 (7·4) | 59 (7·2) | 52 (6·8) | 51 (7·2) |
| Black | 52 (3·9) | 33 (3·4) | 31 (3·8) | 25 (3·3) | 26 (3·7) |
| Other/Mixed | 86 (6·9) | 60 (6·3) | 51 (6·2) | 48 (6·3) | 44 (6·2) |
| **Low Income Household** | | | | | |
| Low income | 125 (11·9) | 91 (11·4) | 85 (12·1) | 74 (11·3) | 72 (12·0) |
| Not low income | 922 (88·1) | 708 (88·6) | 620 (87·9) | 583 (88·7) | 529 (88·0) |
| **Number of Biological Parents in Family** | | | | | |
| Two biological parents | 992 (79·0) | 771 (80·7)* | 659 (79·4) | 629 (81·2) | 584 (81·5) |
| One or no biological parents | 263 (21·0) | 184 (19·3) | 171 (20·6) | 146 (18·8) | 133 (18·5) |
| **Parent Education** | | | | | |
| Highschool or less | 92 (7·3) | 65 (6·8)* | 56 (6·7) | 46 (5·9) | 42 (5·9) |
| Post-secondary diploma or certificate less than Bachelor's degree | 344 (27·4) | 236 (24·7) | 223 (26·8) | 203 (26·1)* | 186 (25·9)* |
| University degree, Bachelor's or above | 820 (65·3) | 654 (68·5) | 552 (66·4) | 528 (68·0) | 489 (68·2) |
| **Urban-rural Residency** | | | | | |
| Large urban centre | 891 (72·3) | 684 (72·5) | 590 (72·0) | 556 (72·8) | 510 (72·0) |
| Small-medium centre | 182 (14·8) | 140 (14·8) | 122 (14·9) | 108 (14·1) | 102 (14·4) |
| Rural area | 159 (12·9) | 120 (12·7) | 107 (13·1) | 100 (13·1) | 96 (13·6) |

*indicates significant (*p* < ·05) baseline predictor of non-response at Times 2–5 in the fully adjusted generalized estimating equations (GEE) models; all values are unweighted.

### Appendix C: Psychometric Analyses of Mental Health Symptom Measure

**Overview and Rationale**

The primary mental health symptom measure used in the ONPATH study is an abbreviated version of the parent-reported Ontario Child Health Study Emotional Behavioural Scales (OCHS-EBS; Duncan et al., 2019). This appendix describes the development, evaluation, and validation of the abbreviated OCHS-EBS measure used in ONPATH.

#### Methods

**Data and Sample**

Psychometric analyses to establish the abbreviated measure used data from the Ontario Child Health Study (OCHS), a provincially representative, cross-sectional survey of 10,802 children and youth aged 4–17 years in Ontario (Boyle et al., 2019b). The OCHS employed a three-stage random cluster sampling design, with data collected via computer-assisted personal interviews and self-completed questionnaires between March and December 2015. A subsample of approximately 700 respondents participated in a test–retest reliability study. The OCHS was selected for measure development and validation for the ONPATH study because it included both the OCHS-EBS and the Mini International Neuropsychiatric Interview for Children and Adolescents (MINI-KID; Sheehand et al., 2010). The MINI-KID served as an external criterion measure and enabled assessment of convergent and discriminant validity of the newly established abbreviated version of the OCHS-EBS for use in the current ONPATH study.

**Measures**

**Original Measure: Parent-Reported Ontario Child Health Study Emotional Behavioural Scales (OCHS:EBS)**

The parent-reported OCHS:EBS (Duncan et al., 2019; Boyle et al., 2019a) is a 52-item questionnaire measuring the following seven DSM-5 disorders in children and youth aged 4 to 17 years: attention-deficit/hyperactivity disorder (ADHD; 8 items), conduct disorder (CD; 11 items), generalized anxiety disorder (GAD; 6 items), major depressive disorder (MDD; 9 items), oppositional defiant disorder (ODD; 6 items), separation anxiety disorder (SAD; 7 items), and social anxiety disorder (social phobia; SP; 5 items). Parents/caregivers are asked to rate symptoms describing each of these disorders in the past 6 months on a scale of 0–2 (0 = "Never or not true", 1 = "Sometimes or somewhat true", 2 = "Often or very true"). Psychometric evaluation using the OCHS data demonstrated adequate item fit for all scales except for conduct disorder and youth-assessed separation anxiety disorder (Duncan et al., 2019).

Previous studies have demonstrated that the OCHS:EBS exhibits good internal and external convergent and discriminant validity for use as a dimensional measure of child and adolescent mental health symptoms consistent with DSM-5 disorders, indicating test-retest reliability estimates over .70 for all scales except for Conduct Disorder (Duncan et al., 2019). Results suggested the parent and youth response for conduct disorder did not meet the empirical standards for reliability and validity.

**Measurement Development and Psychometric Evaluation**

To address ONPATH's research objectives, 29 OCHS-EBS items measuring the following disorders were selected for psychometric evaluation: major depression (9 items), generalized anxiety (6 items), oppositional defiant (6 items) and attention deficit/hyperactivity (8 items). Given low endorsement of conduct disorder symptoms in general population studies (Duncan et al., 2019; Boyle et al., 2019b), coupled with more limited evidence of reliability (Duncan et al., 2025; Duncan et al., 2019; Boyle et al., 2019b), this condition was excluded. Separation and social anxiety were also excluded given pandemic restrictions and more limited opportunities for socialization outside the home.

**External Criterion Measure: Mini International Neuropsychiatric Interview for Children and Adolescents (MINI-KID)**

The MINI-KID is a standardized diagnostic interview for children and youth aged 6–17 years that assesses DSM-IV-TR mental disorders (Sheehand et al., 2010). In the OCHS, the MINI-KID was administered independently to the primary caregiver and youth aged 12 to 17 years and included assessment of past 6-month major depressive episode, generalized anxiety, separation anxiety, social phobia, specific phobia, attention-deficit/hyperactivity disorder, oppositional-defiant disorder, and conduct disorder.

For psychometric validation purposes in the present study, the parent-reported MINI-KID served as an external criterion measure, providing an independent reference for evaluating the convergent and discriminant validity of the abbreviated OCHS-EBS for use in the ONPATH study. To establish comparability across measures, major depressive episode and generalized anxiety disorder were combined—if a child or youth met DSM-IV-TR criteria for either disorder they were classified as having an internalizing disorder. Oppositional defiant disorder and attention-deficit/hyperactivity disorder were each examined separately given that each disorder had comparable measurement across both the abbreviated OCHS-EBS and the MINI-KID. These diagnostic groupings were used to assess the extent to which the abbreviated OCHS-EBS demonstrated expected patterns of association with DSM-IV-TR defined mental disorders.

**Statistical Analyses**

Psychometric analyses of the 29 selected items were conducted using data from the OCHS (Boyle et al., 2019a). In Phase 1, data were randomly partitioned at 50%—resulting in Partition 1 (n = 5,357) and Partition 2 (n = 5,445). An exploratory factor analysis (EFA) on Partition 1 was conducted on all 29 items to determine the dimensions of the scales and identify any potentially problematic items. The determination of the number of factors to retain was based on eigenvalues > 1 and a set of objective model fit indices, including chi-squared statistic, Bayesian Information Criterion (BIC), Comparative Fit Index (CFI), Tucker Lewis Index (TLI) values, and Root Mean Square Error of Approximation (RMSEA). Models with lower chi-squared and BIC values, CFI/TLI > 0·90, and RMSEA < 0·08 were preferred based on the conventional criteria for acceptable model fit (Barrett et al., 2007; Hu & Bentler, 1999).

The performance of each item was then assessed using a series of increasingly restrictive multidimensional item-response-theory (IRT) models. These were based on the graded response model (GRM) for polytomous items (Samejima, 1969) as it accommodates ordered response categories. Two item parameters were estimated for each item: 1) the item *discrimination* parameter and 2) the item *difficulty* parameter. Items with discrimination values below 1·7 and difficulty values beyond the range of -3 to +3 were considered for removal (Baker, 2001, Boatend et al., 2018).

In Phase 2, confirmatory factor analysis (CFA) was conducted on Partition 2, followed by the evaluation of reliability, internal consistency and test-retest reliability with the retained set of items. The internal consistency was assessed via Cronbach's α coefficient with a cutoff of 0·70 for good consistency, while test-retest reliability was evaluated using Pearson's correlation coefficients (acceptable reliability: ≥ 0·70) (Bland & Altman, 1986). Lastly, to assess external convergent and discriminant validity, point-biserial correlation coefficients were used to examine associations between the abbreviated version of the OCHS:EBS for use in the ONPATH study and mental disorder diagnoses ascertained with the MINI-KID. External convergent validity is achieved when correlations are positive between the same constructs across measures (e.g., MINI-KID and OCHS:EBS oppositional defiant symptoms/disorder). External discriminant validity is achieved when correlations of the same constructs between measures (e.g., MINI-KID and OCHS:EBS oppositional defiant symptoms/disorder) are higher than the correlations between different constructs across measures (e.g., MINI-KID attention-deficit/hyperactivity disorder and OCHS:EBS oppositional defiant symptoms) (Cohen & Cohen, 1983).

#### Results

**Phase 1: EFA and IRT-based Analysis**

EFA models with 1 through 5 factors were fitted on Partition 1 under robust maximum likelihood estimation. An oblique rotation was used for solutions with more than one factor. The sample eigenvalues for the first three factors were greater than 1: factor 1 = 8·89, factor 2 = 1·67, factor 3 = 1·01 with the fourth factor eigenvalue below 1 (0·60). Based upon the K1 rule (Kaiser, 1970), where factors are retained with eigenvalues greater than 1, a three-factor model was selected. All objective fit statistics further supported a three-factor model (see Table C1). The model's adequacy was further assessed by examining the parameter estimates, which revealed predominantly high and positive factor loadings on the individual factors (see Table C2 for the factor loadings and factor correlations for the three-factor model). The three factors corresponded to 1) Internalizing, 2) Oppositional Defiant, and 3) Attention Deficit/Hyperactivity Symptom domains. Inter-factor correlations ($r$ = 0·68–0·77) and internal consistency (α = 0·69–0·87) evaluated with Partition 1 provided further support for the three-factor model.

The item '*Deliberately harming self or attempting suicide'* was removed based on EFA results indicating a weak primary loading (factor loadings <0.25) and conceptual concerns because it does not distinguish between self-harm and suicide attempts. Based on the multidimensional graded response IRT model (Table C3), seven additional items were removed according to the pre-specified discrimination parameter criterion (<1·7). As some items may exhibit higher difficulty (threshold) parameters yet hold clinical relevance (e.g., talks about killing self), discrimination parameters were prioritized over difficulty parameters in item removal decisions. This resulted in a 21-item abbreviated version of the OCHS:EBS (retained items are bolded in Table C3): eight internalizing items, six oppositional defiant disorder items, and seven ADHD items. All retained items had discrimination parameters >1·9 and difficulty parameters within the -3 to +3 range, with two exceptions at the highest response category (talks about killing self, b2=4·68; and gets back at people, b2=3·41)

**Phase 2: CFA and Evaluations of Internal Consistency, Test-Retest Reliability, and Internal and External Convergent and Discriminant Validity**

The three-factor model was fitted to Partition 2 using CFA on the retained 21 items, demonstrating excellent model fit (CFI=0·97, TLI=0·97, RMSEA=0·03) and supporting the construct validity of the three-factor structure. Factor loadings are presented in Table C3. Internal convergent and discriminant validity were achieved given the average variance extracted (AVE) values > 0·50 across factors (AVEs = 0·67–0·73). This indicated that the latent constructs explained a substantial proportion of the indicator variance. The three scales demonstrated excellent internal consistency (Cronbach's α = 0·94 to 0·97) and test-retest reliability ($r$ = 0·74 to 0·81).

All correlations between the abbreviated OCHS:EBS scales and corresponding MINI-KID assessments were positive, significant, and ranged from moderate (≥ 0·30) to strong (≥ 0·60) indicating external convergent validity (See Table C4). External discriminant validity was achieved given that all correlations between the abbreviated OCHS:EBS scales and their corresponding MINI-KID assessments were higher than any correlations with the non-corresponding scales.

**Table C1. Model fit statistics of exploratory factor analysis for 29-items of the OCHS:EBS Measure Using Data from the OCHS Partition 1 Sample (n=5,357)**

| | $X^2$ | df | *p* | CFI | TLI | RMSEA | BIC |
|---|---|---|---|---|---|---|---|
| **Factors** | | | | | | | |
| 1 | 3035·44 | 377 | <0·001 | 0·92 | 0·91 | 0·04 | 16067·92 |
| 2 | 1630·54 | 349 | <0·001 | 0·96 | 0·96 | 0·03 | 9936·86 |
| 3 | 943·74 | 322 | <0·001 | 0·98 | 0·98 | 0·02 | 6714·39 |
| 4 | 652·14 | 296 | <0·001 | 0·99 | 0·99 | 0·02 | 5324·50 |
| 5 | 446·97 | 271 | <0·001 | 0·99 | 0·99 | 0·01 | 3770·49 |

**Table C2. Factor loadings from the 3-factor exploratory factor model for 29-items of the OCHS:EBS Measure Using Data from the OCHS Partition 1 Sample (n=5,357)**

| | Factor 1 Internalizing | Factor 2 Oppositional Defiant | Factor 3 Inattention and Hyperactivity |
|---|---|---|---|
| **Items** | | | |
| Unhappy, sad or depressed | **0·52** | 0·41 | 0·12 |
| Has trouble enjoying self | **0·51** | 0·36 | 0·22 |
| Gets no pleasure from usual activities | 0·31 | **0·33** | 0·16 |
| Changes in appetite | 0·28 | **0·29** | 0·14 |
| Trouble sleeping | **0·42** | 0·26 | 0·24 |
| Overtired or lacks energy | **0·45** | 0·32 | 0·09 |
| Feels worthless or inferior | **0·51** | 0·39 | 0·15 |
| Deliberately harms self or attempts suicide | **0·23** | 0·21 | 0·03 |
| Talking about killing self | 0·27 | **0·29** | -0·02 |
| Being too fearful or anxious | **0·65** | 0·05 | 0·27 |
| Worries about doing better at things | **0·46** | 0·17 | 0·05 |
| Finds it hard to stop worrying | **0·71** | 0·11 | 0·14 |
| Anxious or on edge | **0·68** | 0·22 | 0·24 |
| Nervous, high-strung or tense | **0·60** | 0·13 | 0·29 |
| When anxious, his/her mind goes blank | **0·47** | 0·14 | 0·30 |
| Loses temper | 0·19 | **0·59** | 0·33 |
| Argues a lot with adults | 0·16 | **0·57** | 0·30 |
| Blames others for own mistakes | 0·17 | **0·56** | 0·34 |
| Easily annoyed by others | 0·25 | **0·56** | 0·24 |
| Angry and resentful | 0·30 | **0·60** | 0·20 |
| Gets back at people | 0·10 | **0·47** | 0·18 |
| Makes careless mistakes | 0·25 | 0·29 | **0·38** |
| Can't concentrate, can't pay attention for long | 0·23 | 0·12 | **0·69** |
| Fails to finish things he/she starts | 0·21 | 0·29 | **0·56** |
| Distractible, has trouble sticking to any activity | 0·25 | 0·16 | **0·74** |
| Fidgets | 0·19 | 0·17 | **0·66** |
| Can't stay seated when required to do so | 0·09 | 0·14 | **0·68** |
| Impulsive or acts without thinking | 0·19 | 0·39 | **0·56** |
| Has difficulty awaiting turn in games or groups | 0·08 | 0·23 | **0·61** |

Note: The highest factor loadings for each factor are bolded.

**Table C3. Multidimensional IRT results on Partition 1 (29-items; n=5,357) and 3-factor CFA results on Partition 2 for the Abbreviated OCHS:EBS Measure (21-items; n=5,445)**

| | **IRT (29-items) (Partition 1, n=5357)** | | | **3-factor CFA (21-items) (Partition 2, n = 5445)** |
|---|---|---|---|---|
| | **Discrimination** | **Difficulty/Threshold** | | **Factor Loading** |
| **Items** | a | b1 | b2 | |
| *Internalizing Domain* | | | | |
| **Unhappy, sad or depressed** | 2·20 | 0·80 | 3·02 | 0·83 |
| **Has trouble enjoying self** | 2·52 | 1·47 | 2·82 | 0·87 |
| Gets no pleasure from usual activities | 1·62 | 2·04 | 3·85 | ·· |
| Changes in appetite | 1·05 | 1·61 | 4·58 | ·· |
| Trouble sleeping | 1·50 | 1·26 | 3·00 | ·· |
| Overtired or lacks energy | 1·57 | 1·20 | 3·16 | ·· |
| **Feels worthless or inferior** | 2·54 | 1·45 | 2·99 | 0·86 |
| Deliberately harms self or attempts suicide | 1·92 | 2·97 | 4·32 | ·· |
| **Talking about killing self** | 1·98 | 2·72 | 4·68 | 0·86 |
| **Being too fearful or anxious** | 2·38 | 0·92 | 2·46 | 0·85 |
| Worries about doing better at things | 1·42 | 0·51 | 2·91 | ·· |
| **Finds it hard to stop worrying** | 2·83 | 0·94 | 2·29 | 0·77 |
| **Anxious or on edge** | 3·55 | 0·97 | 2·31 | 0·91 |
| **Nervous, high-strung or tense** | 2·54 | 0·85 | 2·47 | 0·88 |
| When anxious, his/her mind goes blank | 1·72 | 1·56 | 3·32 | ·· |
| *Oppositional Defiant Domain* | | | | |
| **Loses temper** | 2·69 | 0·51 | 2·36 | 0·83 |
| **Argues a lot with adults** | 2·47 | 0·97 | 2·42 | 0·79 |
| **Blames others for own mistakes** | 1·93 | 0·54 | 2·50 | 0·79 |
| **Easily annoyed by others** | 2·16 | 0·76 | 2·49 | 0·78 |
| **Angry and resentful** | 2·81 | 1·14 | 2·86 | 0·88 |
| **Gets back at people** | 2·00 | 1·96 | 3·41 | 0·71 |
| *Inattention/Hyperactivity Domain* | | | | |
| Makes careless mistakes | 1·36 | 0·44 | 3·17 | ·· |
| **Can't concentrate, can't pay attention for long** | 2·80 | 0·40 | 1·95 | 0·82 |
| **Fails to finish things he/she starts** | 2·35 | 0·59 | 2·44 | 0·80 |
| **Distractible, has trouble sticking to any activity** | 3·82 | 0·81 | 2·08 | 0·91 |
| **Fidgets** | 2·24 | 0·8 | 2·20 | 0·77 |
| **Can't stay seated when required to do so** | 2·76 | 0·96 | 2·43 | 0·80 |
| **Impulsive or acts without thinking** | 2·32 | 0·78 | 2·38 | 0·87 |
| **Has difficulty awaiting turn in games or groups** | 2·02 | 1·16 | 2·81 | 0·75 |

Note: Bolded = retained items. a = Item difficulty. b1 = Item discrimination for response "Sometimes or Somewhat True". b2 = Item discrimination for response "Often or Very True".

**Table C4. Point-biserial correlation coefficients ($r_{pb}$) between the Abbreviated OCHS:EBS Measure (21-items) and the MINI-KID Using Data from the OCHS Partition 2 Sample (n=5,445)**

| | **OCHS:EBS Internalizing (8 items)** | **OCHS:EBS Oppositional Defiant (6 items)** | **OCHS:EBS Inattention/hyperactivity (7 items)** |
|---|---|---|---|
| MINI-KID Internalizing Disorders | 0·63 | 0·34 | 0·29 |
| MINI-KID Oppositional Defiant Disorder | 0·39 | 0·50 | 0·43 |
| MINI-KID Inattention/Hyperactivity Disorder | 0·31 | 0·36 | 0·50 |

#### Appendix C References


Baker, F. (2001). *The Basics of Item Response Theory*. Washington, DC: ERIC Clearinghouse on Assessment and Evaluation.

Barrett, P. (2007). Structural equation modelling: Adjudging model fit. *Personality and Individual Differences*, *42*(5), 815–824. https://doi.org/10.1016/j.paid.2006.09.018.

Bland, J. M., & Altman, D. (1986). Statistical methods for assessing agreement between two methods of clinical measurement. *The Lancet*, *327*(8476), 307–310. https://doi.org/10.1016/S0140-6736(86)90837-8.

Boateng, G. O., Neilands, T. B., Frongillo, E. A., Melgar-Quiñonez, H. R., & Young, S. L. (2018). Best practices for developing and validating scales for health, social, and behavioral research: a primer. *Frontiers in Public Health*, *6*, 149. https://doi.org/10.3389/fpubh.2018.00149.

Boyle, M. H., Duncan, L., Georgiades, K., Wang, L., Comeau, J., Ferro, M.A., et al. (2019a). The 2014 Ontario Child Health Study Emotional Behavioural Scales (OCHS:EBS) Part II: Psychometric Adequacy for Categorical Measurement of Selected DSM-5 Disorders. *Canadian Journal of Psychiatry, 64(6),* 434–442. https://doi.org/10.1177/0706743718808251.

Boyle, M. H., Georgiades, K., Duncan, L., Comeau, J., Wang, L., & 2014 Ontario Child Health Study Team. (2019b). The 2014 Ontario child health study—methodology. *The Canadian Journal of Psychiatry*, *64*(4), 237–245. https://doi.org/10.1177/0706743719833675.

Cohen, J., & Cohen, P. (1983). Applied Multiple Regression/Correlation Analysis for the Behavioral Sciences. Hillsdale, NJ: Erlbaum.

Duncan, L., Georgiades, K., Wang, L., Comeau, J. & Ferro, M. A et al. (2019). The 2014 Ontario Child Health Study Emotional Behavioural Scales (OCHS:EBS) Part I: A Checklist for Dimensional Measurement of Selected DSM-5 Disorders. *Canadian Journal of Psychiatry, 64(6)*, 423–433. https://doi.org/10.1177/0706743718808250.

Duncan, L., Halladay, J., Bennett, T., Boylan, K., Chang, F., Cometto, J., ... & Lipman, E. (2025). Psychometric evaluation of the Ontario Child Health Study Emotional Behavioural Scales (OCHS-EBS) in children and youth from an outpatient mental health setting. *Psychiatry Research*, *354*, 116809. https://doi.org/10.1016/j.psychres.2025.116809.

Hu, L. T., & Bentler, P. M. (1999). Cutoff criteria for fit indexes in covariance structure analysis: Conventional criteria versus new alternatives. *Structural Equation Modeling: A Multidisciplinary Journal*, *6*(1), 1–55. https://doi.org/10.1080/10705519909540118.

Kaiser HF. 1970. A second-generation little jiffy. *Psychometrika*, *35*, 401–415. https://doi.org/10.1007/BF02291817.

Sheehan, D. V., Sheehan, K. H., Shytle, R. D., Janavs, J., Bannon, Y., Rogers, J. E., ... & Wilkinson, B. (2010). Reliability and validity of the mini international neuropsychiatric interview for children and adolescents (MINI-KID). *The Journal of Clinical Psychiatry*, *71*(3), 17393. https://doi.org/10.4088/jcp.09m05305whi.

#### Appendix D: Derivation of Mental Health Symptom Domain Score based on Moderated Non-Linear Factor Analyses

To establish the outcome scores for the primary longitudinal analysis, we applied moderated nonlinear factor analysis (MNLFA) on the 2023 longitudinal ONPATH sample (n = 1261) to 1) evaluate measurement invariance across time and various participant characteristics and 2) derive scores adjusted for non-invariance on the 21-item OCHS:EBS Abbreviated Measure. The MNLFA is a novel and more flexible approach to testing measurement invariance that allows for the simultaneous evaluation of multiple sources of respondent bias, including continuous covariates (Bauer, 2017). Upon confirming that the assumption of unidimensionality of each OCHS:EBS domain was met by testing with single-factor CFA, we applied MNLFA to identify the items with differential item functioning (DIF) related to child and family characteristics (see Figure D1 for the model specification). To identify meaningful covariates for further MNLFA, we performed a logistic-regression-based DIF detection test (Choi, Gibbons, & Crane, 2011) with McFadden's pseudo $R^2 > 0{\cdot}013$ as a cutoff for flagging items with potential DIF (Zumbo, 1999). If any item was flagged with DIF by a certain covariate, that covariate will be included in the MNLFA model. As a result, we included child's age (continuous; range=3·2–20·7 years), polynomial term of child's age (continuous/mean-centered; for capturing nonlinear age effects), child's gender (categorical; 0 = girl, 1 = boy), parent's gender (categorical; 0 = woman, 1 = man), parent's racialized identity (categorical; 0 = non-White, 1 = White), and parent distress (continuous/mean-centered; range = 0–24). These variables were also selected based on our concern surrounding the broad age range of our child/youth participants, and previous literature about proxy respondent biases related to sex at birth, gender identity, race and ethnic identity, and parent distress/psychopathology in mental health assessments (Olino et al., 2020; Stevanovic et al., 2017; Vitoratou et al., 2019). Data collection method (online vs. phone) was not included in the MNLFA given its ignorable DIF effects across all items (all McFadden's pseudo $R^2 < .009$).

Given the longitudinal nature of our data, two calibration samples of independent observations (n = 1261 each) were pseudo-randomly drawn from the full sample of repeated observations (n = 4538) to ensure similar distribution of child's age across samples for testing model stability of DIF identification. Following the steps described in the prior literature (Gottfredson et al., 2019; Sifre et al., 2021), the DIF identification process started with a baseline model by allowing the latent mean and variance regressed on all the covariates, while excluding the polynomial (i.e., age$^2$) effect on latent variance for computation efficiency as previously suggested (Gottfredson et al., 2019). A criterion of $p < 0{\cdot}10$ was used to retain effects of the covariates on the latent mean and variance in the baseline model. Next, covariate effects were introduced on the item intercept and factor loading for one item at a time while treating other items as anchor items (i.e., items without DIF). This process was repeated for each of the items under the each of our three latent mental health domain factors with a $p < 0{\cdot}05$ cut-off for retaining covariate effects on the item intercept and factor loading in the DIF model. Finally, all covariates with effects that met the alpha criteria in the previous steps across the calibration samples were included simultaneously with the full sample. In the final model, only covariates with effects that survived the $p < 0{\cdot}05$ criterion were retained for obtaining the latent factor estimates. All the MNLFA steps were performed separately for each of the three OCHS:EBS domains under the unidimensionality assumption. Once the MNLFA process was complete, we extracted the derived factor scores adjusted for any detected measurement non-invariance. The adjusted factor scores were then used as the main outcome variables for the longitudinal analysis.

As a result, the unidimensionality assumption of the 3 mental health domains (Internalizing Symptoms, Oppositional Defiant Symptoms, Inattention/Hyperactivity Symptoms) was confirmed with CFA using the data collected in the ONPATH study (Table D1). The final MNLFA model with the retained covariate effects on latent factor mean/variance and item intercept/loading for each OCHS:EBS-SF21 mental health domain is presented in Table D2. Child's age was the strongest moderator of item functioning across domains given that 17 out of 21 items were flagged with age-related DIF ($|t| = 2{\cdot}50$ to $9{\cdot}38$, all $p < 0{\cdot}05$). The items "*difficulty awaiting turn in games or groups*" showed the largest age-related intercept DIF ($t = -9{\cdot}38$, $p < 0{\cdot}001$), indicating that when holding the latent trait (i.e., level of inattention/hyperactivity symptoms) constant, parents of younger children were more likely to endorse this item. As for child's gender, parents of boys tended to endorse some oppositional defiant and inattention/hyperactivity items (e.g., *"blames others for own mistakes"*) more frequently when controlling for the symptom level ($t = 2{\cdot}64$ to $6{\cdot}09$, all $p < 0{\cdot}05$), while parents of girls were more likely to endorse items such as "*easily annoyed by others*" more frequently when controlling for the symptom level ($t = -4{\cdot}89$, $p < 0{\cdot}001$).

Parent's characteristics also impacted the evaluation of their child's behaviour ($|t| = 2{\cdot}04$ to $5{\cdot}16$, all $p < 0{\cdot}05$). For instance, parents who identified as non-racialized, White were more likely to endorse some oppositional defiant and inattention/hyperactivity items (e.g., "*angry and resentful*" and "*can't stay seated when required to do so*"), while racialized parents were more likely to endorse the internalizing behaviour item "*unhappy, sad or depressed*". Parents who identified as a woman tended to endorse internalizing and oppositional defiant items (e.g., "*being too fearful/anxious*"; $t = -2{\cdot}58$ to $-5{\cdot}96$, all $p < 0{\cdot}05$) while the opposite holds for some oppositional defiant and inattention/hyperactivity items (e.g., "*difficulty awaiting turn in games or groups*"; $t = 3{\cdot}62$ to $4{\cdot}24$, all $p < 0{\cdot}001$). Parent distress showed overall smaller moderating effects with its largest effect observed in the oppositional defiant item "*angry and resentful*" ($t = 5{\cdot}10$, $p < 0{\cdot}001$), which was more likely to be endorsed by parents with more distress. Overall, the loading DIF effects were minimal and mostly related to age ($|t| = 2·19$ to $5·00$, all $p < 0·05$). For instance, "*loses temper*" and "*blames others for own mistakes*" are more associated with the latent factor of oppositional defiant symptoms in older children.

After accounting for these item-level biases, larger effects of age and parent distress compared to other covariates on latent means were observed across domains. Specifically, a concave downward effect of age was observed for internalizing symptoms (DIF by age $t = 8·40$ & by age$^2$ $t = -6·00$, both $p < 0·001$) while lower oppositional defiant and inattention/hyperactivity symptoms were associated with older age ($t = -9·71$ & $-4·83$, both $p < 0·001$). Children of parents with more distress tended to have higher scores across the three domains ($t = 16·20$ to $21·00$, all $p < 0·001$). On average, girls had higher internalizing symptoms ($t = -7·77$, $p < 0·001$) while boys had higher inattention/hyperactivity symptoms ($t = 6·60$, $p < 0·001$), but no gender difference was found for oppositional defiant symptoms. Children of non-racialized, White parents tended to have higher scores across domains ($t = 2·76$ to $3·07$, all $p < 0·01$). Also, parents who identified as a woman tended to report higher internalizing and inattention/hyperactivity symptoms of their child ($t = -3·86$ & $-2·00$, $p < 0·05$). Further, the variations of the latent factor scores (i.e., latent variance) across domains increased with child's age and level of parent distress ($t = 5·00$ to $8·17$, all $p < 0·001$).

Descriptive statistics of MNLFA-derived factor scores are presented in Table D3. The MNLFA scores were highly correlated with raw sum scores across domains ($r = 0·97$ to $0·98$), but the former was more normally distributed (see Figure D2) and provided invariant estimates across waves and participant characteristics for a more accurate estimation of growth trajectories in the primary longitudinal analyses.

**Table D1. Model fit statistics of one-factor confirmatory factor analysis for the test of unidimensionality on the ONPATH sample**

| | T1 (n=1260) | T2 (n=958) | T3 (n=831) | T4 (n=778) | T5 (n=711) |
|---|---|---|---|---|---|
| **Internalizing** | | | | | |
| CFI | 0·99 | 0·98 | 0·99 | 0·99 | 0·99 |
| TLI | 0·98 | 0·97 | 0·98 | 0·99 | 0·98 |
| RMSEA | 0·08<br>[0·07, 0·09] | 0·10<br>[0·09, 0·12] | 0·08<br>[0·07, 0·10] | 0·07<br>[0·06, 0·08] | 0·09<br>[0·07, 0·10] |
| Factor loading range | 0·70–0·91 | 0·69–0·88 | 0·756–0·897 | 0·72–0·91 | 0·70–0·90 |
| **Oppositional Defiant** | | | | | |
| CFI | 0·99 | 0·99 | 0·99 | 0·99 | 0·99 |
| TLI | 0·99 | 0·99 | 0·99 | 0·99 | 0·99 |
| RMSEA | 0·05<br>[0·04, 0·07] | 0·06<br>[0·04, 0·08] | 0·05<br>[0·02, 0·07] | 0·06<br>[0·04, 0·08] | 0·06<br>[0·04, 0·08] |
| Factor loading range | 0·62–0·85 | 0·67–0·85 | 0·72–0·86 | 0·66–0·88 | 0·73–0·84 |
| **Inattention/hyperactivity** | | | | | |
| CFI | 0·98 | 0·98 | 0·98 | 0·98 | 0·99 |
| TLI | 0·97 | 0·97 | 0·97 | 0·96 | 0·98 |
| RMSEA | 0·10<br>[0·08, 0·11] | 0·10<br>[0·09, 0·12] | 0·10<br>[0·08, 0·11] | 0·11<br>[0·10, 0·13] | 0·07<br>[0·06, 0·09] |
| Factor loading range | 0·63–0·90 | 0·70–0·91 | 0·60–0·92 | 0·69–0·90 | 0·67–0·89 |

**Figure D1. MNLFA model specification**

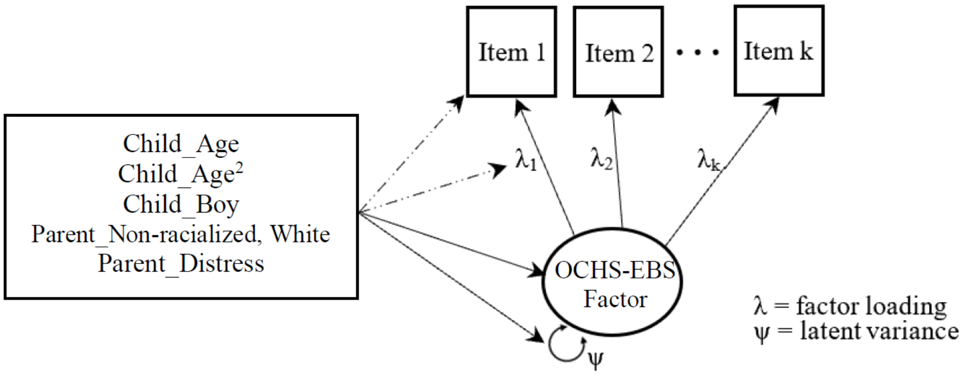

***Table D2*. *Final MNLFA Model (beta coefficients and standard errors of retained DIF effects).***

| | Child_Age | | Child_Age$^2$ | | Child_Boy | | Parent_Man | | Parent_Non-Racialized (White) | | Parent Distress | |
|---|---|---|---|---|---|---|---|---|---|---|---|---|
| **Items** | ***int*** | ***load*** | ***int*** | ***load*** | ***int*** | ***load*** | ***int*** | ***load*** | ***int*** | ***load*** | ***int*** | ***load*** |
| **Internalizing** | | | | | | | | | | | | |
| Unhappy, sad or depressed | ·· | 0·06 (0·01)*** | ·· | ·· | ·· | ·· | ·· | ·· | 0·29 (0·10)** | ·· | ·· | ·· |
| Trouble enjoying self | 0·03 (0·01)** | ·· | ·· | ·· | ·· | ·· | ·· | ·· | ·· | ·· | ·· | ·· |
| Feeling worthless or inferior | -0·01 (0·01) | 0·05 (0·02)** | -0·01 (0·00)** | ·· | ·· | ·· | -0·45 (0·09)*** | ·· | ·· | ·· | ·· | ·· |
| Talking about killing self | ·· | ·· | ·· | ·· | ·· | ·· | ·· | ·· | ·· | ·· | ·· | ·· |
| Being too fearful or anxious | -0·06 (0·02)*** | ·· | 0·01 (0·00)*** | ·· | -0·24 (0·10)* | ·· | -0·60 (0·10)*** | ·· | ·· | ·· | ·· | ·· |
| Finding it hard to stop worrying | 0·04 (0·01)* | -0·04 (0·02)* | ·· | ·· | ·· | ·· | -0·30 (0·10)** | ·· | ·· | ·· | -0·03 (0·01)** | ·· |
| Anxious or on edge | 0·01 (0·01) | ·· | 0·02 (0·00)*** | ·· | ·· | ·· | ·· | ·· | ·· | ·· | ·· | ·· |
| Being nervous, high-strung or tense | ·· | ·· | ·· | ·· | ·· | ·· | ·· | ·· | ·· | ·· | ·· | ·· |
| Latent Mean | 0·04 (0·01)*** | ·· | -0·01 (0·00)*** | ·· | -0·27 (0·04)*** | ·· | -0·14 (0·04)*** | ·· | 0·14 (0·04)** | ·· | 0·11 (0·01)*** | ·· |
| Latent Variance | 0·05 (0·01)*** | ·· | ·· | ·· | ·· | ·· | ·· | ·· | ·· | ·· | 0·04 (0·01)*** | ·· |
| **Oppositional Defiant** | | | | | | | | | | | | |
| Loses temper | -0·09 (0·02)*** | 0·05 (0·02)** | ·· | ·· | ·· | ·· | ·· | ·· | ·· | ·· | ·· | ·· |
| Argues a lot with adults | -0·05 (0·01)*** | ·· | -0·01 (0·00)*** | ·· | 0·22 (0·08)** | ·· | ·· | ·· | ·· | ·· | ·· | ·· |
| Blames others for own mistakes | -0·05 (0·01)*** | 0·03 (0·01)** | -0·01 (0·00)*** | ·· | 0·49 (0·08)*** | ·· | -0·220 (0·08)** | ·· | ·· | ·· | ·· | ·· |
| Easily annoyed by others | 0·11 (0·01)*** | ·· | -0·01 (0·00)*** | ·· | -0·41 (0·08)*** | ·· | -0·22 (0·09)* | ·· | ·· | -0·35 (0·13)** | 0·05 (0·01)*** | ·· |
| Angry and resentful | ·· | ·· | ·· | ·· | ·· | ·· | -0·25 (0·09)** | ·· | ·· | ·· | 0·05 (0·01)*** | ·· |
| Gets back at people | -0·09 (0·02)*** | ·· | -0·02 (0·00)*** | ·· | 0·38 (0·09)*** | ·· | 0·34 (0·10)*** | ·· | -0·39 (0·12)** | ·· | ·· | ·· |
| Latent Mean | -0·03 (0·01)*** | ·· | 0·00 (0·00) | ·· | -0·03 (0·04) | ·· | -0·07 (0·04) | ·· | 0·14 (0·05)** | ·· | 0·08 (0·01)*** | ·· |
| Latent Variance | 0·05 (0·01)*** | ·· | ·· | ·· | ·· | ·· | ·· | ·· | ·· | ·· | 0·05 (0·01)*** | ·· |
| **Inattention/Hyperactivity** | | | | | | | | | | | | |
| Can’t concentrate or pay attention for long | 0·14 (0·02)*** | ·· | ·· | ·· | -0·32 (0·13)* | 0·38 (0·13)** | ·· | ·· | ·· | ·· | ·· | -0·04 (0·01)** |
| Fails to finish things he/she starts | 0·04 (0·01)** | 0·06 (0·01)*** | ·· | ·· | ·· | ·· | ·· | ·· | ·· | ·· | ·· | ·· |
| Distractible/trouble sticking to any activity | 0·10 (0·02)*** | ·· | ·· | ·· | -0·39 (0·15)* | 0·37 (0·15)* | ·· | ·· | ·· | ·· | -0·04 (0·01)** | ·· |
| Fidgets | ·· | ·· | ·· | ·· | ·· | ·· | ·· | ·· | 0·18 (0·09)* | ·· | 0·03 (0·01)** | ·· |
| Can’t stay seated when required to do so | -0·11 (0·02)*** | -0·04 (0·02)* | ·· | ·· | 0·41 (0·10)*** | ·· | ·· | ·· | -0·58 (0·11)*** | ·· | ·· | ·· |
| Impulsive or acts without thinking | -0·04 (0·01)*** | ·· | ·· | ·· | 0·27 (0·07)*** | ·· | ·· | ·· | ·· | ·· | ·· | ·· |
| Difficulty awaiting turn in games or groups | -0·12 (0·01)*** | ·· | 0·01 (0·00)** | ·· | ·· | 0·24 (0·08)** | 0·35 (0·08)*** | ·· | ·· | -0·30 (0·12)* | 0·04 (0·01)*** | ·· |
| Latent Mean | -0·07 (0·01)*** | ·· | -0·00 (0·00)** | ·· | 0·26 (0·04)*** | ·· | -0·07 (0·03)* | ·· | 0·12 (0·05)** | ·· | 0·08 (0·01)*** | ·· |
| Latent Variance | 0·04 (0·01)*** | ·· | ·· | ·· | ·· | ·· | ·· | ·· | ·· | ·· | 0·04 (0·01)*** | ·· |

Note. int=item intercept; load=factor loading; $^{*}p < 0·05$, $^{**}p < 0·01$, $^{***}p < 0·001$; Int: The probability of endorsing an item is dependent upon the covariate, when holding constant where someone is positioned on the latent variable. (e.g., a positive intercept DIF effect by age means that older children are more likely to endorse the categories that indicate higher symptoms for that item); Load: The association between the item and the latent variable is weaker or stronger dependent upon the covariate (e.g., a positive loading DIF effect by age means that the item was more highly associated with the latent factor for older children); Latent mean: the latent factor scores vary by covariates (e.g., a positive age effect on latent mean indicates that older children tend to have higher factor scores); Latent variance: the variance of latent factor scores varies by covariates (e.g., a positive age effect on latent variance indicates older children have more heterogeneous factor scores).

**Table D3. Descriptive statistics of MNLFA vs. raw sum scores derived from the 21-item OCHS:EBS Abbreviated Measure**

| | Internalizing | | | | | Oppositional Defiant | | | | | Inattention/Hyperactivity | | | | |
|---|---|---|---|---|---|---|---|---|---|---|---|---|---|---|---|
| | *T1* | *T2* | *T3* | *T4* | *T5* | *T1* | *T2* | *T3* | *T4* | *T5* | *T1* | *T2* | *T3* | *T4* | *T5* |
| **MNLFA[a] Scores** | | | | | | | | | | | | | | | |
| N | 1260 | 958 | 831 | 778 | 711 | 1260 | 958 | 830 | 778 | 712 | 1260 | 958 | 831 | 778 | 712 |
| Mean | -0·14 | -0·12 | -0·21 | -0·29 | -0·29 | 0·17 | 0·22 | 0·00 | -0·09 | -0·12 | 0·24 | 0·36 | 0·06 | 0·00 | -0·02 |
| SD | 10·07 | 10·04 | 1·06 | 1·07 | 1·04 | 0·98 | 0·99 | 0·95 | 0·97 | 0·94 | 1·03 | 1·06 | 0·98 | 1·03 | 1·02 |
| Skewness | 0·51 | 0·44 | 0·53 | 0·60 | 0·61 | 0·35 | 0·33 | 0·48 | 0·58 | 0·52 | 0·24 | 0·13 | 0·28 | 0·41 | 0·37 |
| Kurtosis | -0·28 | -0·29 | -0·08 | -0·29 | -0·04 | -0·02 | -0·07 | 0·17 | 0·12 | 0·08 | -0·48 | -0·45 | -0·37 | -0·23 | -0·30 |
| **Raw Sum Scores[b]** | | | | | | | | | | | | | | | |
| N | 1247 | 949 | 819 | 768 | 700 | 1251 | 956 | 826 | 772 | 707 | 1250 | 949 | 822 | 766 | 696 |
| Mean | 4·04 | 4·12 | 3·77 | 3·53 | 3·48 | 3·67 | 3·85 | 3·19 | 2·98 | 2·92 | 4·53 | 4·98 | 3·99 | 3·85 | 3·76 |
| SD | 3·71 | 3·61 | 3·60 | 3·64 | 3·50 | 2·80 | 2·86 | 2·69 | 2·70 | 2·64 | 3·51 | 3·66 | 3·29 | 3·45 | 3·36 |
| Skewness | 0·88 | 0·82 | 0·97 | 1·04 | 1·05 | 0·63 | 0·61 | 0·82 | 0·95 | 0·92 | 0·58 | 0·46 | 0·70 | 0·84 | 0·83 |
| Kurtosis | 0·02 | -0·01 | 0·37 | 0·33 | 0·44 | -0·12 | -0·09 | 0·30 | 0·50 | 0·40 | -0·50 | -0·65 | -0·14 | 0·00 | -0·07 |

a: MNLFA scores were latent factor scores derived from the model specified in Figure D1 for each respective subscale. Missing items were handled using full information maximum likelihood (FIML) estimation.
b: Raw sum scores were calculated by summing internalizing, oppositional defiant, and inattention/hyperactivity subscale items (pro-rated up to 25% missing items, with mean score of completed items being multiplied by total number of items for pro-ration).

**Figure D2. Distribution of raw sum scores and MNLFA scores for ONPATH samp**

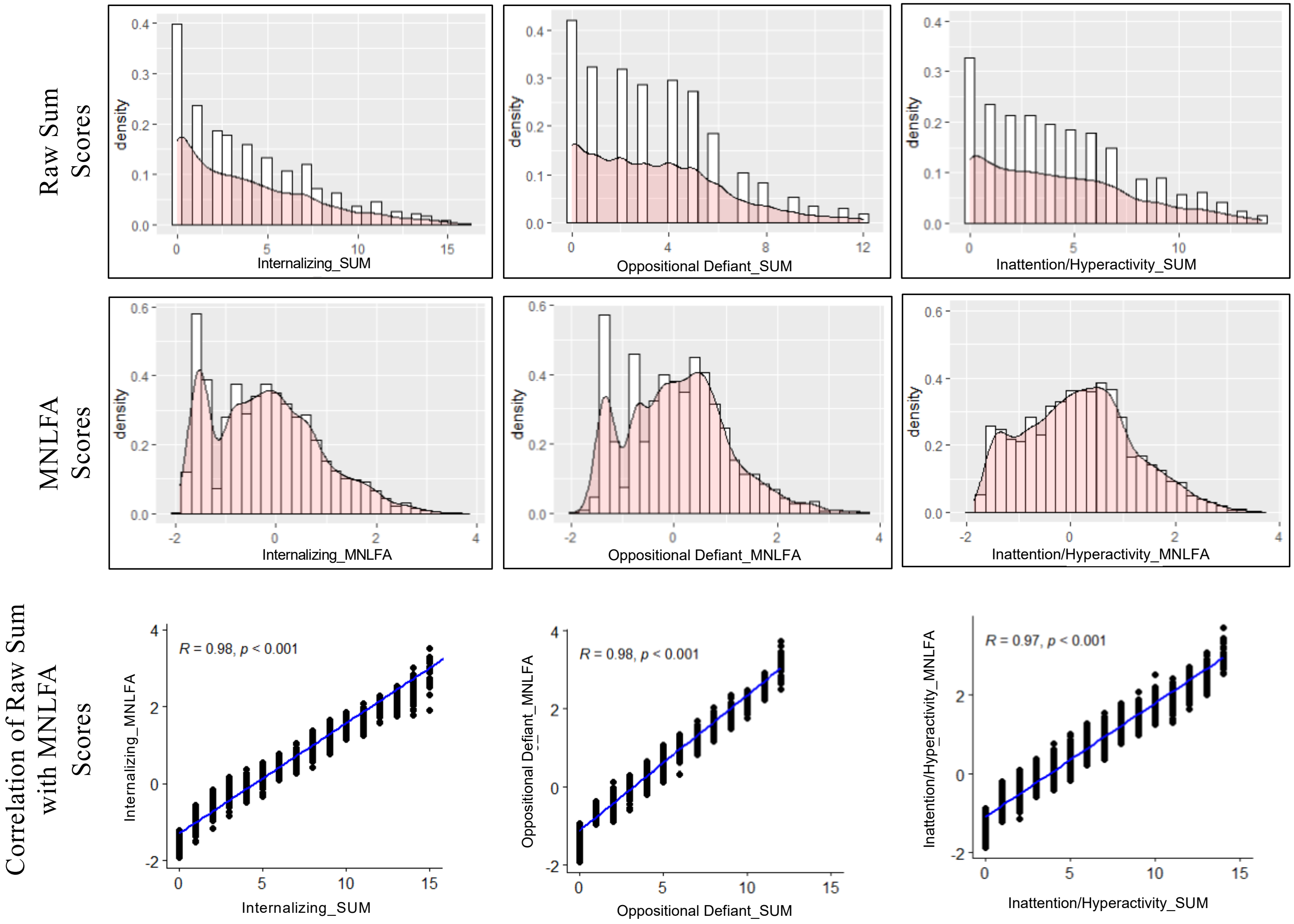

#### Appendix D References


Bauer, D. J. (2017). A more general model for testing measurement invariance and differential item functioning. *Psychological methods*, *22*(3), 507. https://psycnet.apa.org/doi/10.1037/met0000077.

Choi, S. W., Gibbons, L. E., & Crane, P. K. (2011). Lordif: An R package for detecting differential item functioning using iterative hybrid ordinal logistic regression/item response theory and Monte Carlo simulations. *Journal of statistical software*, *39*, 1-30. https://doi.org/10.18637/jss.v039.i08.

Cole, V. T., Gottfredson, N. C., Giordano, M., & Janssen, T. (2021). Automated fitting of moderated nonlinear factor analysis (MNLFA) through the Mplus program. *R package version*, *1*(0).

Gottfredson, N. C., Cole, V. T., Giordano, M. L., Bauer, D. J., Hussong, A. M., & Ennett, S. T. (2019). Simplifying the implementation of modern scale scoring methods with an automated R package: Automated moderated nonlinear factor analysis (aMNLFA). *Addictive behaviors*, *94*, 65-73. https://doi.org/10.1016/j.addbeh.2018.10.031.

Olino, T. M., Guerra-Guzman, K., Hayden, E. P., & Klein, D. N. (2020). Evaluating maternal psychopathology biases in reports of child temperament: An investigation of measurement invariance. *Psychological assessment*, *32*(11), 1037. https://doi.org/10.1037/pas0000945.

Stevanovic, D., Jafari, P., Knez, R., Franic, T., Atilola, O., Davidovic, N., ... & Lakic, A. (2017). Can we really use available scales for child and adolescent psychopathology across cultures? A systematic review of cross-cultural measurement invariance data. *Transcultural Psychiatry*, *54*(1), 125-152. https://doi.org/10.1177/1363461516689215.

Vitoratou, S., Garcia-Rosales, A., Banaschewski, T., Sonuga-Barke, E., Buitelaar, J., Oades, R. D., ... & Chen, W. (2019). Is the endorsement of the Attention Deficit Hyperactivity Disorder symptom criteria ratings influenced by informant assessment, gender, age, and co-occurring disorders? A measurement invariance study. *International Journal of Methods in Psychiatric Research*, *28*(4), e1794. https://doi.org/10.1002/mpr.1794.

Zumbo, B. D. (1999). A handbook on the theory and methods of differential item functioning (DIF). *Ottawa: National Defense Headquarters*, *160*, 53.

#### Appendix E: Item Non-response for Study Variables

**Table E1. Missing Data for Time Invariant Main Study Variables at T1, Valid Cases (n) and Percent Missing**

| | Baseline Valid n | % Missing |
|---|---|---|
| Child Gender | 1,238 | 1·8 |
| Child Age | 1,261 | 0·0 |
| Pandemic Lockdown Status | 1,237 | 1·9 |
| Pre-Existing Clinician Diagnosed Physical Mental or Neurodevelopmental Conditions | 1,250 | 0·9 |
| Parent Gender | 1,255 | 0·5 |
| Parent Racialized Identity | 1,241 | 1·6 |
| Low Income Household | 1,047 | 17·0 |
| Number of Biological Parents in Home | 1,255 | 0·5 |

All values are unweighted.

**Table E2. Missing Data for Time Variant Variables by Time of Data Collection, Valid Cases (n) and Percent Missing**

| | Baseline Valid n | % Missing | T2 Valid n | % Missing | T3 Valid n | % Missing | T4 Valid n | % Missing | T5 Valid n | % Missing |
|---|---|---|---|---|---|---|---|---|---|---|
| All | 1,261 | 0·0 | 959 | 0·0 | 833 | 0·0 | 779 | 0·0 | 719 | 0·0 |
| Schooling Modality | 1,210 | 0·2 | 915 | 0·2 | 800 | 0·2 | 736 | 0·3 | 675 | 0·3 |
| COVID-19 Case Rate per 100 000* | 1,237 | 1·9 | 948 | 1·2 | 823 | 1·2 | 760 | 2·4 | 711 | 1·1 |
| Parental Distress** | 1,256 | 0·4 | 954 | 0·5 | 830 | 0·4 | 779 | 0·0 | 718 | 0·1 |
| Internalizing Symptoms** | 1,247 | 1·1 | 947 | 1·3 | 823 | 1·2 | 768 | 1·4 | 700 | 2·6 |
| Oppositional Defiant Symptoms** | 1,251 | 0·8 | 951 | 0·8 | 827 | 0·7 | 772 | 0·9 | 707 | 1·7 |
| Inattention/Hyperactivity Symptoms** | 1,250 | 0·9 | 950 | 0·9 | 828 | 0·6 | 766 | 1·7 | 696 | 3·2 |

All values are unweighted, *Missing due to missing FSA **Missing on the scale level.

#### Appendix F: Supplementary Results for the Primary Analysis

**Table F1. Model fit statistics of latent growth curve model (LGCM) in different functional forms**

| Model | $\chi^2$ | df | CFI | TLI | RMSEA [95% CI] |
|---|---|---|---|---|---|
| **Internalizing** | | | | | |
| Linear | 17·47 | 10 | 0·99 | 0·99 | 0·02 [0·00, 0·04] |
| Quadratic | 13·21 | 6 | 0·99 | 0·99 | 0·03 [0·01, 0·05] |
| Piecewise (T2 knot)* | 15·48 | 8 | 0·99 | 0·99 | 0·03 [0·00, 0·05] |
| **Oppositional Defiant** | | | | | |
| Linear | 48·25 | 10 | 0·972 | 0·97 | 0·06 [0·04, 0·07] |
| Quadratic | 37·05 | 6 | 0·977 | 0·96 | 0·06 [0·05, 0·09] |
| Piecewise (T2 knot)* | 40·80 | 8 | 0·976 | 0·97 | 0·06 [0·04, 0·08] |
| **Inattention/ Hyperactivity** | | | | | |
| Linear | 57·99 | 10 | 0·97 | 0·97 | 0·06 [0·05,0 ·08] |
| Quadratic | 48·93 | 6 | 0·97 | 0·95 | 0·08 [0·06, 0·10] |
| Piecewise (T2 knot)* | 54·50 | 8 | 0·97 | 0·96 | 0·07 [0·05, 0·09] |

*Covariances between the first piece of the slope parameter and the other two growth parameters (intercept and the second slope) were fixed to 0 for the purpose of model identification.

**Figure F1. LGCM with covariates**

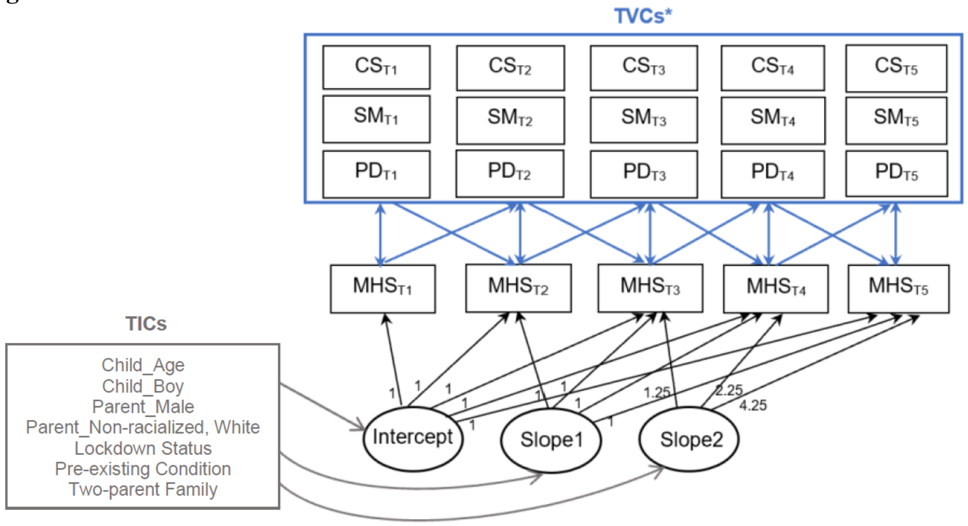


Note. MHS=mental health symptoms (internalizing, oppositional defiant, and inattention/hyperactivity symptoms were modeled separately); TICs=time-invariant covariates; TVCs=time-varying covariates; CS=COVID-19 case rate; SM=schooling mode; PD= parental distress. *The three TVCs were simultaneously specified with covariances between the repeated measures, which were not depicted in the plot.
